# Utilizing Electronic Health Records (EHR) and Tumor Panel Sequencing to Demystify Prognosis of Cancer of Unknown Primary (CUP) patients

**DOI:** 10.1101/2022.12.22.22283696

**Authors:** Intae Moon, Jaclyn LoPiccolo, Sylvan C. Baca, Lynette M. Sholl, Kenneth L. Kehl, Michael J. Hassett, David Liu, Deborah Schrag, Alexander Gusev

## Abstract

When a standardized diagnostic test fails to locate the primary site of a metastatic cancer, it is diagnosed as a cancer of unknown primary (CUP). CUPs account for 3-5% of all cancers but do not have established targeted therapies, leading to typically dismal outcomes. Here, we develop OncoNPC, a machine learning classifier of CUP, trained on targeted next generation sequencing data from 34,567 tumors across 22 primary cancer types collected as part of routine clinical care at three institutions under AACR Project GENIE initiative [1]. OncoNPC achieved a weighted F1 score of 0.94 for high confidence predictions on known cancer types (65% of held-out samples). To evaluate its clinical utility, we applied OncoNPC to 971 CUP tumor samples from patients treated at the Dana-Farber Cancer Institute (DFCI). OncoNPC CUP subtypes exhibited significantly different survival outcomes, and identified potentially actionable molecular alterations in 23% of tumors. Importantly, patients with CUP, who received first palliative intent treatments concordant with their OncoNPC predicted sites, showed significantly better outcomes (Hazard Ratio 0.348, 95% C.I. 0.210 - 0.570, p-value 2.32×10^−5^) after accounting for potential measured confounders. As validation, we showed that OncoNPC CUP subtypes exhibited significantly higher polygenic germline risk for the predicted cancer type. OncoNPC thus provides evidence of distinct CUP subtypes and offers the potential for clinical decision support for managing patients with CUP.

## Introduction

When a standardized diagnostic work-up, including radiology and pathology review, fails to locate the primary site of a metastatic cancer, it is diagnosed as a cancer of unknown primary (CUP). CUP represents about 3-5% of all cancers worldwide [2] and is characterized by aggressive progression and poor prognosis (survival of 6 to 16 months [3]). The hidden nature of the primary cancer types for a CUP limits treatment options since clinical responses to some treatments are known to vary based on patients’ tumor types (e.g., identical BRAF V600 mutations targetable in melanoma but no colorectal cancer[4]). Emerging cancer treatments targeting actionable molecular alterations are typically developed for specific cancer types: HER2 in breast cancer and EGFR mutation or ALK/ROS1 rearrangement in Non-small cell lung cancer (NSCLC) [5], and are thus inaccessible to CUP patients. Accurately identifying the latent primary site for CUPs and demonstrating clinical benefit from site-specific therapies may thus open many existing treatment options for patients with CUP.

Pathology review plays a key role in determining primary cancer types of malignant tumors based on immunohistochemistry (IHC) results as well as tumor morphology and clinical findings; however, pathological diagnosis can be challenging for highly metastatic or poorly differentiated tumors. For known cancer types, prior studies showed that an IHC-based diagnostic work-up correctly identified 77 - 86% of primary tumors, which further decreased to 60 - 71% for metastatic tumors [6]. For patients with CUP, IHC results suggestive of a single primary diagnosis account for only 25% of tumors [3]. The subjective nature of pathological interpretation and guidelines, as well as the variability in IHC staining techniques across institutions thus makes it challenging to establish consistent protocols for CUP diagnosis [7].

Molecular tumor profiling has been proposed as an alternative for CUP primary classification due to its quantitative nature and high accuracy on tumors with known cancer types [8–12]. Such tools rely on microarray DNA methylation [8], whole genome sequencing (WGS) [9, 12], or RNA-seq data [11] to train machine learning classifiers using reference data from known-primary tumors. However, molecular sequencing remains prohibitive and not integrated into the existing standard of care, limiting the translational potential of such methods. Recently, key work by Penson et al. [10] demonstrated that accurate primary cancer type classifications could be made from next generation sequencing (NGS) of targeted panels, now routinely collected at many cancer centers and applicable to hundreds of thousands of tumors [1]. However, its clinical utility in diagnosing and aiding treatment for patients with CUP was not systematically investigated.

Several recent studies have investigated the potential clinical benefit of molecular CUP classification, in non-randomized prospective studies [13–15] as well as the randomized clinical trials [16]. These trials have often struggled to recruit sufficient numbers of representative patients and explore the full range of available therapies. A recent randomized phase II trial [16] did not find significant improvement in 1-year survival for the treatment group receiving site-specific therapy guided by molecular profiling. However, this study was limited by a small number of patients (n = 101) recruited over 7 years, with few common solid tumor types and well-established therapies [17]. Assessing the clinical benefits of molecular CUP classification thus poses both an opportunity for precision medicine and a major challenge for conventional randomized studies.

In contrast to prospective trials, retrospective Electronic Health Records (EHR) data can capture a larger and more heterogeneous patient population, despite potential biases due to informative missingness and unobserved heterogeneity. Coupling EHR data with tumor sequencing can offer insights into the molecular mechanisms of CUPs and their relationship to clinical outcomes. As panel sequencing is often part of the standard of care, such insights also have the potential to assist diagnostic efforts and clinical management within existing molecular workflows. Here, we utilized multi-center, Next Generation Sequencing (NGS) targeted panel sequencing data from 36,445 tumor samples with known primary cancers to train and evaluate a machine learning classifier predicting a primary cancer type of a given tumor sample. We applied this classifier, named *OncoNPC* (**Onco**logy **N**GS-based **P**rimary cancer type **C**lassifier), to 971 patients with CUP with clinical follow up at the Dana-Farber Cancer Institute (DFCI). Using the OncoNPC cancer type predictions, we identified CUP subtypes that shared specific characteristics with their corresponding predicted primaries including: significant differences in clinical outcomes, elevated germline risk, and prognostic somatic alterations. 23% of OncoNPC classified CUP tumors had actionable somatic variants enabled by their corresponding OncoNPC cancer type predictions. Finally, using EHR-based treatment and survival data, we showed that site-specific treatments concordant with the OncoNPC cancer type predictions led to longer survival than those discordant with the cancer type predictions. Our findings suggest that many CUPs can be classified into meaningful subtypes with the potential to aid clinical decision making.

## Results

### OncoNPC accurately classifies 22 known cancer types

We developed *OncoNPC* (**Onco**logy **N**GS-based **P**rimary cancer type **C**lassifier), a molecular cancer type classifier trained on multicenter targeted panel sequencing data (Fig. 1). OncoNPC utilized all somatic alterations including mutations (single nucleotide variants and indels), mutational signatures, copy number alterations, as well as patient age at sequencing and sex to jointly predict cancer type using a XGBoost algorithm (see Methods). Importantly, no other aspects of tumor morphology, pathology, or patient demographics were used so as not to bias the classifier towards known cancers. OncoNPC was trained and validated on the processed data consisting of 29,176 primary and metastasis tumor samples from 22 known cancer types collected at the DFCI, MSK, and VICC (see Table 1 for details). Across all 22 cancer types, OncoNPC achieved a weighted F1 score of 0.784 on the held-out test tumor samples consisting of 7,289 tumor samples (weighted precision and recall : 0.789 and 0.791, respectively). Across 10 cancer groups (grouped by sites and treatment options (Table 1), OncoNPC achieved an overall weighted F1 score of 0.824 (weighted precision and recall : 0.829 and 0.826, respectively). Despite the evident class imbalance across cancer types, OncoNPC showed well-balanced precision across the cancer types (Fig. 2a) and cancer groups (Fig. 2b). Thresholding on prediction confidence (*p*_*max*_, the maximum posterior probability across all labels) further increased the performance: weighted F1 score of 0.830 with 91.6 % remaining samples at *p*_*max*_ *≥* 0.5 and 0.942 with 65.2 % remaining samples at *p*_*max*_ *≥* 0.9 (Fig. 2c, 2d). While rarer cancer types had generally lower overall performance, increasing the *p*_*max*_ threshold reduced this difference between common/rare cancer types (Fig. 2c, 2d). At *p*_*max*_ *≥* 0, common cancer types in the upper quartile in terms of the number of tumor samples (NSCLC, BRCA, COADREAD, DIFG, PRAD, and PAAD) had a mean F1 of 0.84 while rare cancer types in the lower quartile (WDTC, MNGT, GINET, PANET, AML, and NHL) had a mean F1 of 0.58, whereas at *p*_*max*_ *≥* 0.9 common and rare cancer had a mean F1 of 0.95 and 0.86, respectively. This demonstrates that the OncoNPC was still able to do high-quality predictions for a subset of tumor samples in rare cancer types, for which training data was limited.

**Table 1.**
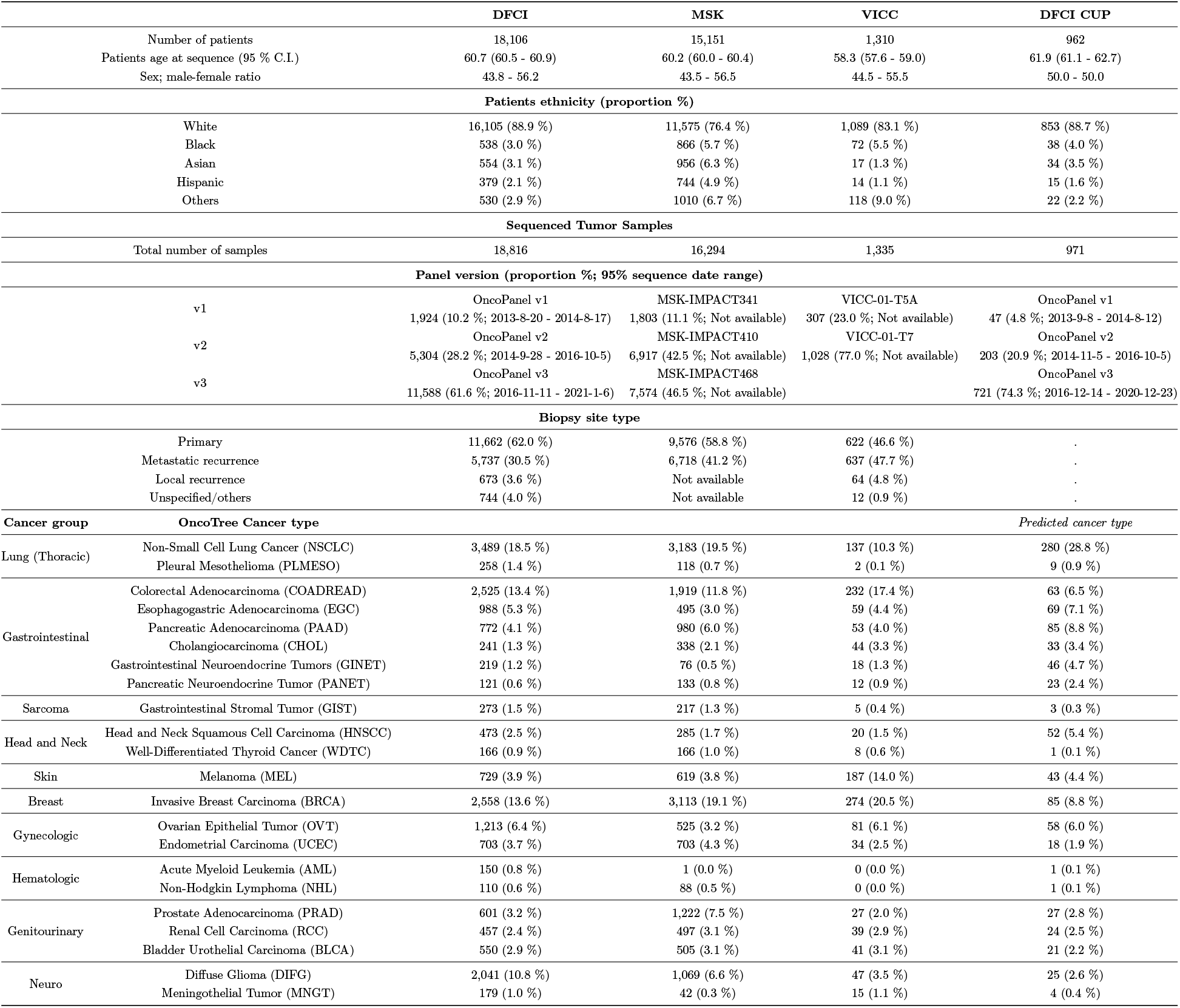
Demographic details of the patients and tumor samples across DFCI, MSK, and VICC.

**Figure 1.**
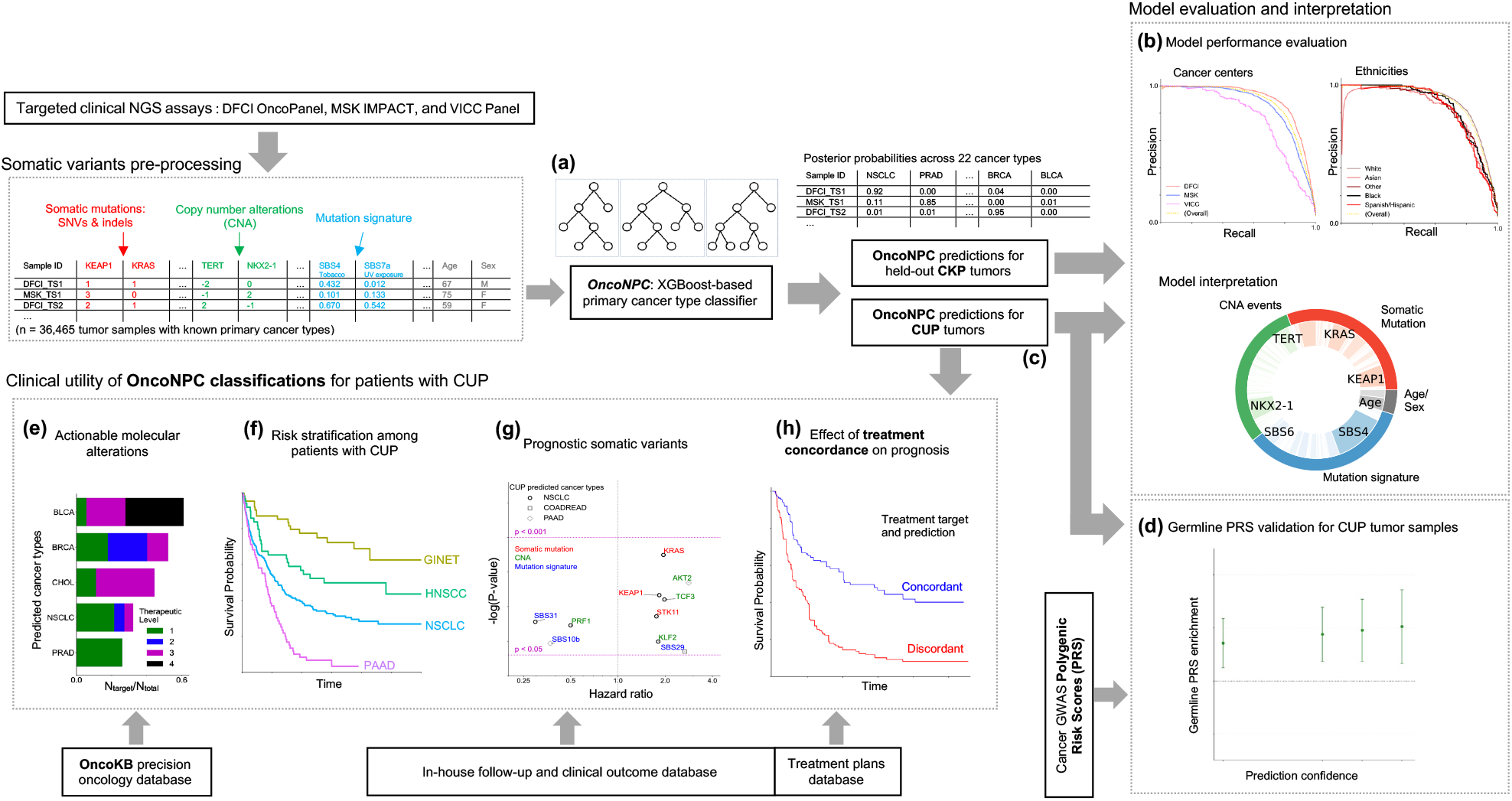
Overview of model development and analysis workflow. **(a)** OncoNPC, a XGBoost-based classifier, was trained and evaluated using 36,729 tumor samples across 22 cancer types from Cancers of Known Primary (CKP) collected from three different cancer centers. **(b)** OncoNPC performance was evaluated on the held-out tumor samples (n = 7,289). **(c)** OncoNPC was applied to 971 CUPs at a single institution to predict primary cancer types. OncoNPC predicted CUP subtypes were then investigated for association with: **(d)** elevated germline risk, **(e)** actionable molecular alterations, **(f)** overall survival, and **(g)** prognostic somatic features. **(h)** A subset of CUP patients with detailed treatment data were evaluated for treatment-specific outcomes.

**Figure 2.**
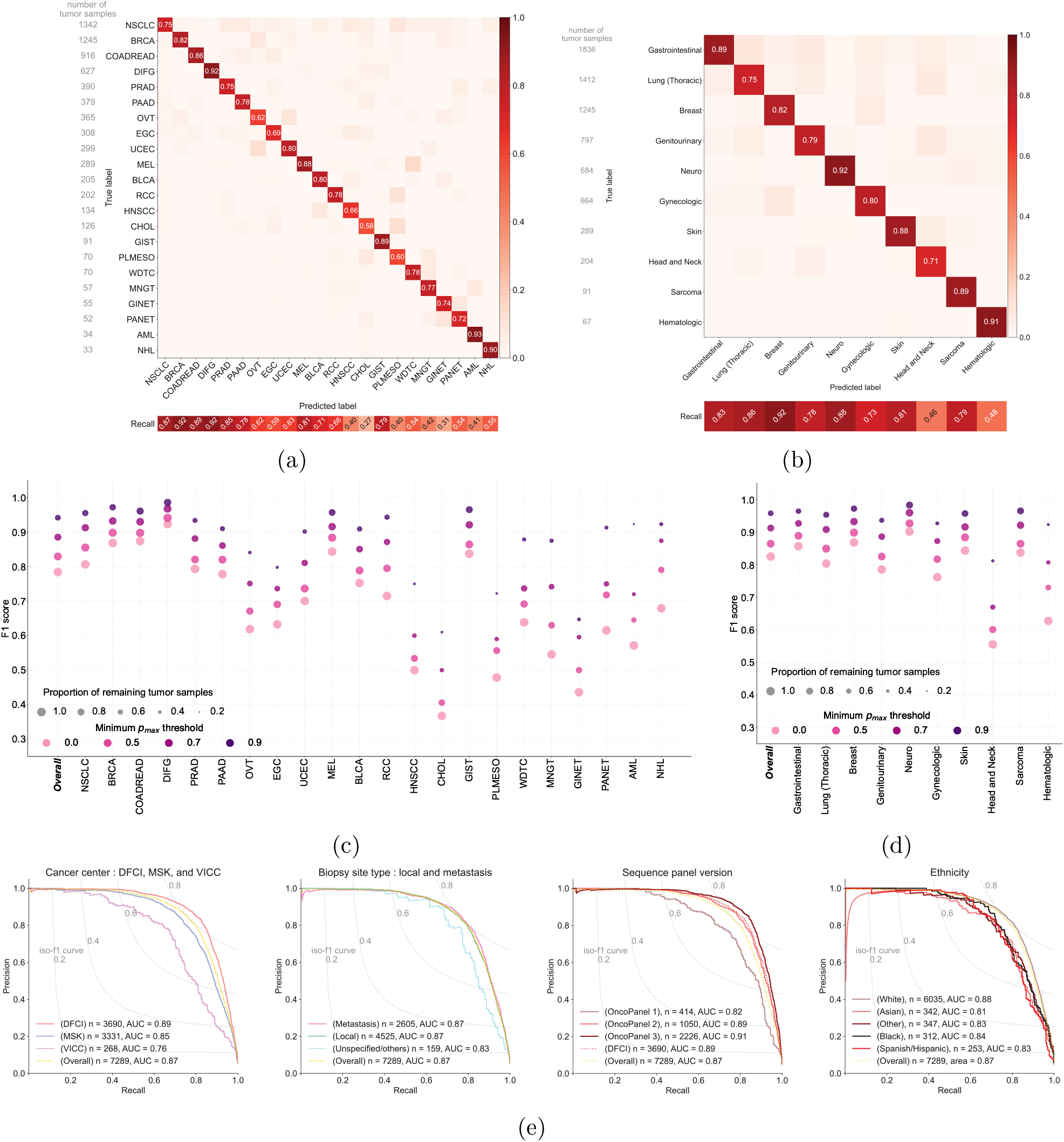
Cancer type prediction performance of OncoNPC. **(a)**,**(b)** The normalized confusion matrix of OncoNPC classification performance on the held-out test set (n = 7,289) for **(a)** 22 detailed cancer types and **(b)** 10 broad cancer groups based on site and treatment (see Table 1). The sensitivity for each cancer type or cancer group is shown below each confusion matrix and the sample size is shown to the left of each confusion matrix. **(c), (d)** The performance (by F1 score) of OncoNPC on the test set across cancer types **(c)** and groups **(d)** at 4 different prediction confidences (i.e., minimum *p*_*max*_ thresholds). Each dot size is scaled by the proportion of tumor samples retained. **(e)** Precision-recall curves showing the performance of the OncoNPC across different cohorts in the test set by: cancer center, biopsy site type, sequence panel version, and ethnicity (color coded), with the yellow dotted curve corresponding to the baseline performance on the full test set.

OncoNPC achieved robust performance against potential dataset shifts due to the factors including cancer center, biopsy site type, sequence panel version, and patient ethnicity (Fig. 2e). OncoNPC showed comparable performance for tumor samples from DFCI (AUC-PR, area under the precision recall curve = 0.89, n = 3,690) and those from MSK (AUC-PR = 0.85, n = 3,331). OncoNPC performance for those from VICC was slightly lower (AUC-PR = 0.76, n = 268). OncoNPC showed comparable performance for primary tumor samples (AUC-PR = 0.87, n = 4,525) and metastatic tumor samples (AUC-PR = 0.87, n = 2,605), demonstrating its capability to predict the primary cancer site of metastatic cancers without loss of performance. To assess the OncoNPC performance over time, we investigated its performance across sequence panel versions utilized at DFCI, as the panel version is a proxy for sequence dates of tumor samples (see Table 1). The OncoNPC performance on tumor samples from earlier versions of DFCI sequence panels (OncoPanel v1 : AUC-PR = 0.82, n = 414 and OncoPnael v2 : AUC-PR = 0.89, n = 1,050) was slightly lower than the performance on the tumor samples from the most recent panel (OncoPanel v3 : AUC-PR = 0.91, n = 2,226) which also contained the largest number of genes. As all tumor samples have been collected from OncoPanel v3 since October 2016, we expect our model to make high-quality predictions in a prospective setting. Finally, OncoNPC demonstrated consistent performance across patient ethnicity, an important consideration to avoid introducing algorithmic disparities. See Supplementary Fig. S1a for more detailed center-specific OncoNPC performance.

### Applying OncoNPC to CUP tumor samples

We applied OncoNPC to classify 971 CUP tumors from patients who were admitted to DFCI and sequenced as part of routine clinical care. Compared to the held-out cohort of Cancer with Known Primary (CKP; n = 7,289), OncoNPC classifications for CUPs had prediction probabilities lower than those of the DFCI held-out cohort of Cancer with Known Primary (CKP; n = 3,690), but comparable to those of the DFCI held-out cohort of CKPs including other cancer types (n = 8,025), indicating that CUPs may contain other hard-to-classify cancer types: mean prediction probability 0.764 (95% C.I. 0.750 - 778) for CUPs versus 0.881 (95% C.I. 0.875 - 0.887) for the held-out CKPs at DFCI and 0.769 (95% C.I. 0.764 - 0.774) for all held-out CKPs at DFCI (Fig. S1 and Supplementary Fig. S1b). However, more than half of the CUP tumors (518/971) could still be classified with high confidence (i.e., prediction probability > 0.8), and multiple classified types had distributions of posterior probabilities comparable to their corresponding CKPs: Non-small Cell Lung Cancer (NSCLC), Invasive Breast Carcinoma (BRCA), Pancreatic Adenocarcinoma (PAAD), Prostate Adenocarcinoma (PRAD), and Gastrointestinal Neuroendocrine Tumors (GINET). Interestingly, CUPs with predicted GINET were highly confident, despite their small number of tumor samples in the training cohort (n = 359; 0.99% of the training cohort), suggesting some rarer cancer types may nevertheless be confidently identifiable. As shown in Fig. 3b, the most common CUP cancer types were Non-small Cell Lung Cancer (NSCLC), Pancreatic Adenocarcinoma (PAAD), Invasive Breast Carcinoma (BRCA), Esophagogastric Adenocarcinoma (EGC), and Colorectal Adenocarcinoma (COADREAD); of which NSCLC, BRCA, and COADREAD were also the most common CKP types. These rates are broadly consistent with prior findings that the most frequently revealed underlying primary cancers for CUPs by autopsy include lung, large bowel, and pancreas cancers [18]. Finally, comparable rates were observed upon applying OncoNPC to 581 CUP tumors at MSK (Supplementary Fig. S4)

**Figure 3.**
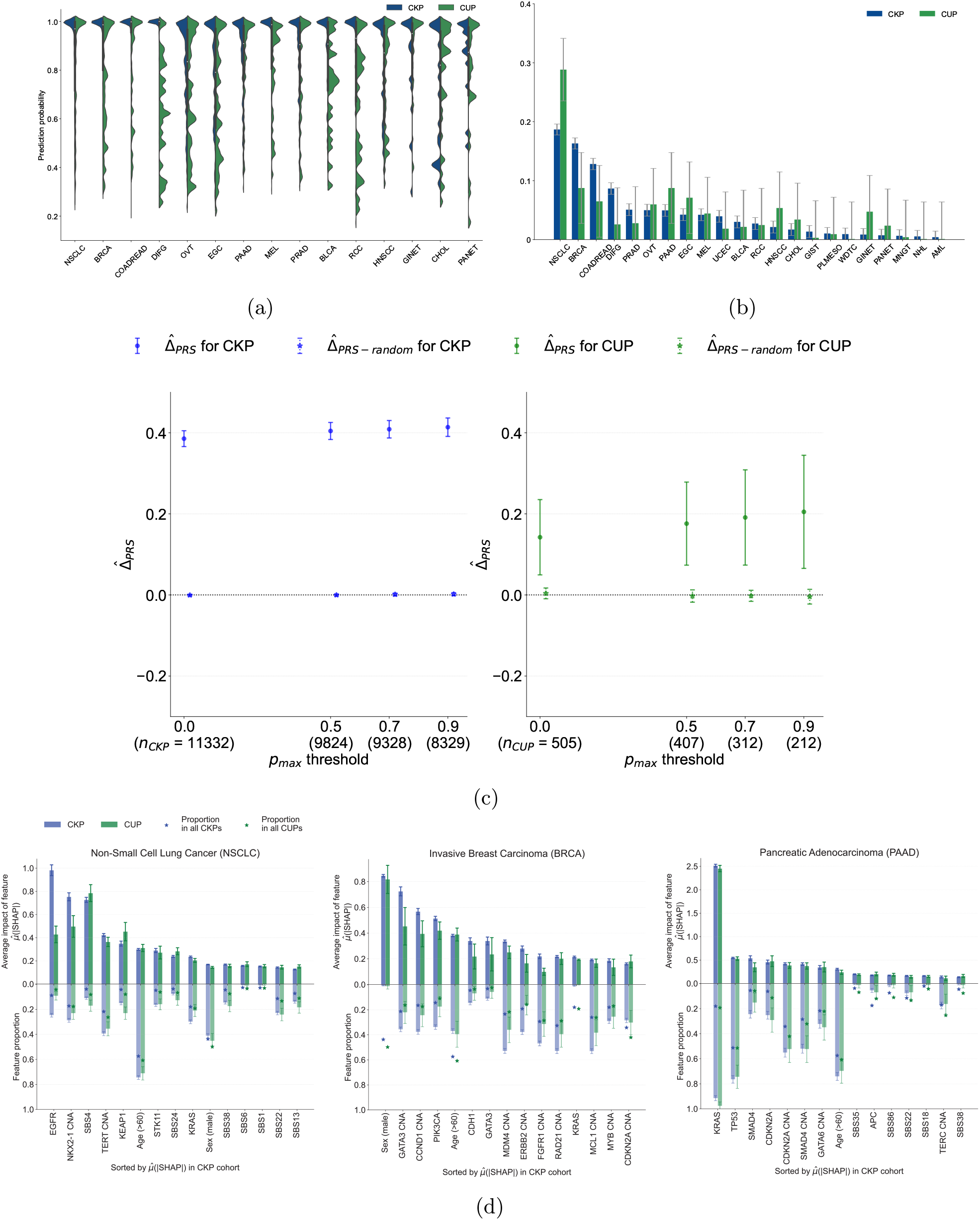
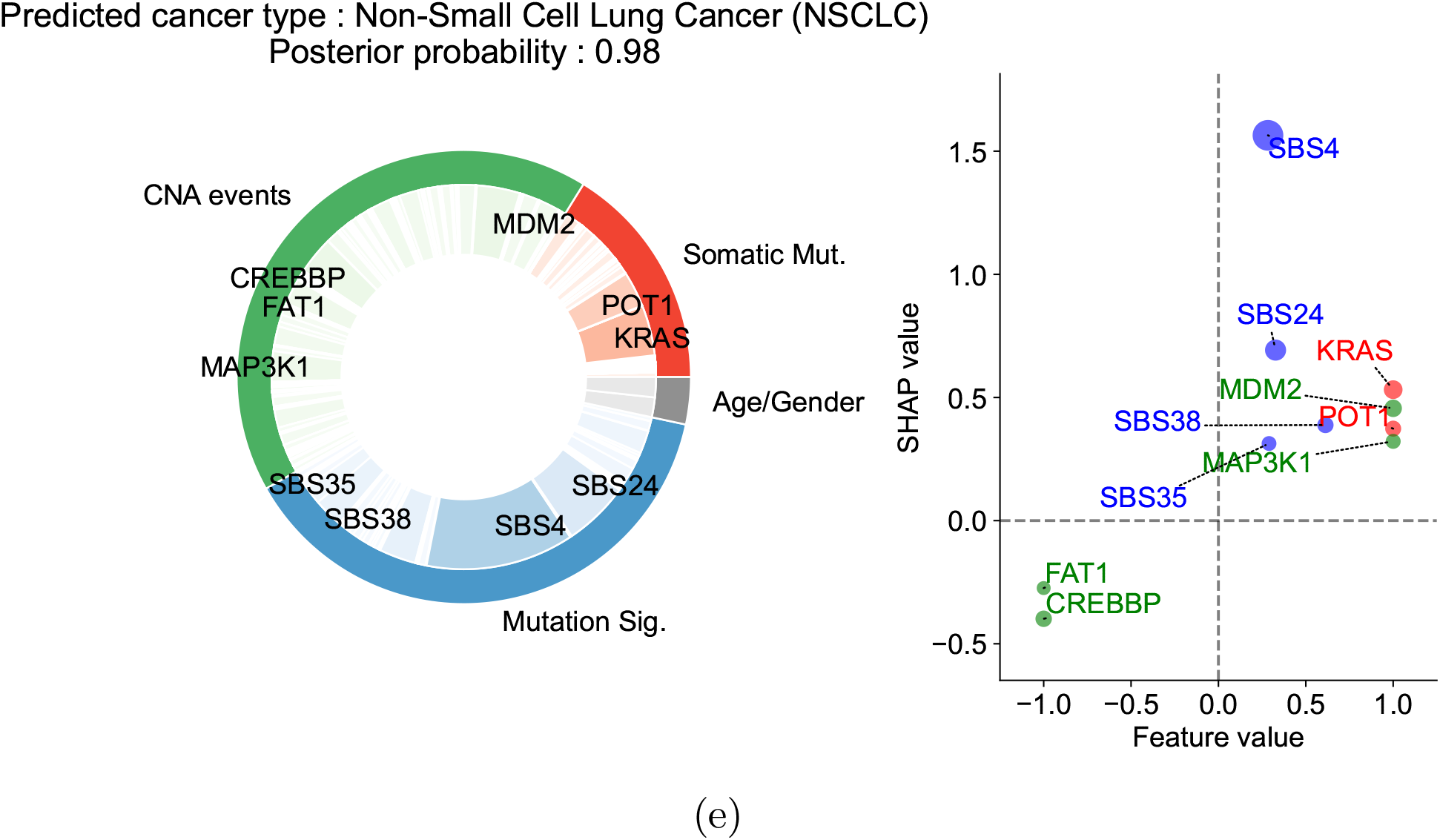
Applying OncoNPC to CUP tumor samples and interpreting cancer type predictions. **(a)** Empirical distributions of prediction probabilities for correctly predicted, held-out CKP tumor samples (n = 3,429) and CUP tumor samples (n = 934) across CKP cancer types (blue) and their corresponding OncoNPC predicted cancer types for CUP tumors (green). Only OncoNPC classifications with at least 20 CUP tumor samples are shown. **(b)** Proportion of each CKP cancer type and the corresponding OncoNPC predicted CUP cancer type. All training CKP tumor samples (n = 36,445) and all held-out CUP tumor samples (n = 971) are shown. For both **(a)** and **(b)**, the cancer types (x-axis) are ordered by the number of CKP tumor samples in each cancer type. **(c)** Germline Polygenic Risk Score (PRS) enrichment of the CKP tumor samples (n = 11,332) and CUP tumor samples with available PRS data (n = 505) averaged across 8 cancer types. The magnitude of the enrichment is quantified by 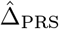: the mean difference between the concordant (i.e. OncoNPC matching) cancer type PRS and mean of PRSs of discordant cancer types (see Methods). 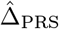 is shown for CKPs in blue (for reference) and CUPs in green. As a negative control, 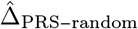 is also shown after permuting the OncoNPC labels. **(d)** Top 15 most important features based on mean absolute SHAP values (i.e., 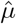 [19]) for the top 3 most frequent cancer types in the cohort: Non-Small Cell Lung Cancer (NSCLC), Invasive Breast Carcinoma (BRCA), and Pancreatic Adenocarcinoma (PAAD). The carrier rate for each feature in corresponding CKP and CUP cancer cohorts as well as the entire CKP and CUP cohorts are shown as bars going downwards and star-shaped markers, respectively. For mutation signature features that have continuous values, individuals with feature values one standard deviation above the mean were treated as positives and the rest as negative. For age, individuals above the population mean were treated as positives and the rest as negatives. **(e)** Explanation of OncoNPC cancer type prediction for a sample patient with CUP. The patient is a 76 year-old male, with a tumor biopsy from the liver. The pie chart on the left shows the Top 10 important features across three different feature categories (i.e., CNA events, somatic mutation, and mutation signatures), and the scatter plot on the right shows their SHAP values and feature values. The size of each dot is scaled by corresponding absolute SHAP value.

### Explaining OncoNPC cancer type predictions

OncoNPC learns complex non-linear relationships between input somatic variants and clinical features and provides interpretable primary cancer type predictions, where impact of each input feature on a prediction is quantified as a SHAP value [19]. We investigated the most impactful features in predicting each cancer type across the CKP and CUP cohorts to evaluate face validity of OncoNPC (see Fig. 3d for the top 3 most frequent cancer types in the cohort: NSCLC, BRCA, and PAAD, and Supplementary Fig. S2 and S3 for other cancer types). For NSCLC, the most important features were EGFR mutation and SBS4, a tobacco smoking-associated mutation signature [20], for CKP tumor samples and CUP with NSCLC predicted tumor samples, respectively; both consistent with the known etiology of lung cancer. Somatic mutation in the EGFR gene is frequently observed in NSCLC tumors and the gene itself is a well-known therapeutic target for patients with NSCLC [21, 22]. Carcinogens in tobacco smoke have been known to cause lung cancer [23]. For BRCA, the most important feature for both CKP and CUP was sex, as expected, followed by CNA events in GATA3 and CCND1 genes, known drivers and prognostic indicators in breast cancer [24, 25]. For PAAD, KRAS mutation was significantly more common than the population averages and by far the most important somatic feature. Mutations in the KRAS gene occur frequently among patients with colorectal cancer and are known to have prognostic significance [26, 27].

OncoNPC provides intuitive illustrations of an explanation for individual-level predictions (Fig. 3e). As an example, we show the explained classification for a tumor sample biopsied from the liver of the 76 year-old male patient and subsequently diagnosed with CUP. From the chart review, we found that the patient reported a 60-pack year smoking history, as well as having lived near a tar and chemical factory as a child. Despite the CUP diagnosis, OncoNPC confidently classified the primary site as NSCLC with posterior probability of 0.98. SBS4, a tobacco smoking-associated mutation signature, was significantly enriched in the patient’s tumor sample, which has, by far, the most impact on the prediction; followed by SBS24 mutation signature associated with known exposures to aflatoxin [20]; and KRAS mutation. Note that inhalation of aflatoxin has been linked to cause primary lung cancer [28–30], and KRAS mutation is one of the most common drivers of NSCLC [31, 32]. The feature interpretation analysis demonstrated that OncoNPC was able to capture biologically consistent, cancer-type specific signals from interpretable somatic mutation and clinical features at an individual tumor level as well as a cohort level.

### Germline PRS-based validation on CUP tumor samples

We hypothesized that, if OncoNPC was accurately identifying latent primary cancers, the classified CUP cancer types would exhibit increased germline risk for the corresponding cancers. To that end, we imputed common germline variation for each CUP patient and quantified their polygenic risk scores (PRS) across 8 common cancers using external cancer GWAS data (see Methods). PRSs are a continuous estimate of the underlying germline liability for a given cancer and orthogonal from the somatic data used to train OncoNPC. As hypothesized, patients with CUP had a significantly higher mean germline PRS for the OncoNPC predicted cancers (Fig. 3c and see Supplementary Fig. S6 for cancer type-specific analysis) compared to other cancer types. The magnitude of the difference (i.e., 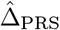) increased for more confident OncoNPC predictions (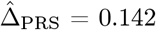, 95% C.I. 0.0494 – 0.235, Wald test p-value: 2.66 × 10^−3^ and 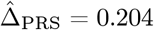, 95% C.I. 0.0655 – 0.344, Wald test p-value: 3.98 × 10^−3^ at *p*_max_ threshold = 0.0 and *p*_max_ threshold = 0.9, respectively). As a negative control, the same analysis conducted with randomly shuffled OncoNPC labels showed no enrichment. As a positive control, the same analysis conducted on CKPs, with available imputed PRS (n = 11,332), also demonstrated a highly significant germline enrichment, as expected. Notably, the enrichment for CUPs was in between that of CKPs and random tumors, suggesting that while OncoNPC classified CUPs are genetically correlated with CKPs, they still exhibit additional heterogeneity.

### OncoNPC-based risk stratification among patients with CUP

To demonstrate clinical utility of OncoNPC, we examined if OncoNPC cancer type predictions can stratify risk among patients with CUP. Using overall survival, we identified subtypes which had significant prognostic differences in median survival based on the OncoNPC classifications (Figure 4a, Chi-squared test, p-value: 4.90 × 10^−14^). Overall, the poorest prognosis was observed in patients with CUP predicted to be Esophagogastric Adenocarcinoma (EGC) and Pancreatic Adenocarcinoma (PAAD): median survival 8.44 months for the combined cohort (95% C.I. 5.39 - 10.5, n = 107). The most favorable prognosis was observed in patients with CUP predicted to be Head and Neck Squamous Cell Carcinoma (HNSCC), Gastrointestinal Neuroendocrine Tumors (GINET), and Pancreatic Neuroendocrine Tumors (PANET): median survival 48.2 months for HNSCC (95% C.I. 19.6 - not estimable, n = 41) and not estimable median survival (i.e. the estimated survival curve never reached the median) for the combined GINET and PANET cohort (n = 57), respectively. Our identified favorable subtypes are consistent with established favorable CUP subtypes such as poorly or well differentiated neuroendocrine carcinomas of unknown primary and squamous cell carcinoma of non-supraclavicular cervical lymph nodes [33]. OncoNPC subtypes can thus be leveraged to meaningfully stratify patients by expected median survival.

**Figure 4.**
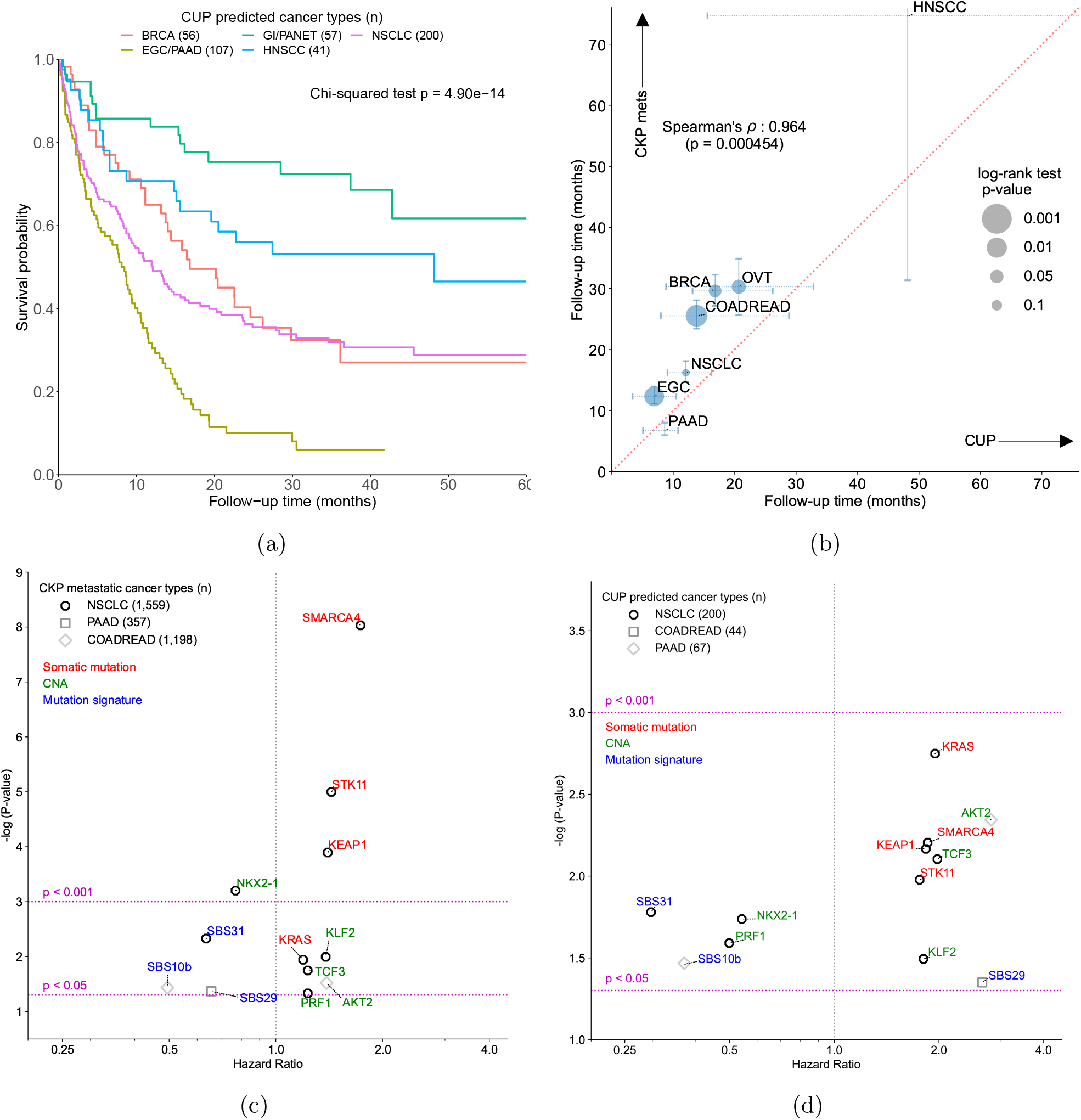
Consistent survival and prognostic biomarkers between OncoNPC classifications and known cancers. **(a)** Survival stratification for patients with CUP based on their OncoNPC predicted cancer types. The Kaplan-Meier estimator [64] was used to estimate survival probability for each predicted cancer type over the follow-up time of 60 months from sequence date, with statistical significance assessed by Chi-square test. **(b)** Correspondence between median survival time (in months) of CUP predicted cancer types (x-axis) and those of metastatic CKP cancer types (y-axis): Spearman’s rho 0.964 (p-value: 4.54 × 10^− 4^). The size of each dot reflects the p-value of log-rank test for significant difference in median survival between CUP - metastatic CKP pairs. Only cancer types with at least 30 CUP tumor samples having OncoNPC probabilities greater than 0.5 are shown. **(c), (d)** Prognostic somatic variants significantly associated with overall survival, shared between three different CUP **(c)**-metastatic CKP **(d)** pairs (NSCLC, PAAD, and COAD-READ; indicated by point shape). Variant types are indicated by colors: red for somatic mutations, green for CNAs, and blue for mutation signatures.

### CUP-CKP metastatic survival comparison

We investigated if cancer-specific prognosis is shared between CUP predicted cancer and their corresponding CKP metastatic cancers. Utilizing overall survival data linked to the National Death Index and in-house follow-up data (see Methods), we found that median survival times of CUP-metastatic CKP pairs were significantly correlated across the cancer types (Spearman’s *ρ*: 0.964, p-value: 4.54 × 10^− 4^; Fig. 4b). This significant relationship provides evidence that genetics-based OncoNPC predictions capture prognostic signals specific to each predicted cancer type. While correlated, median survival times were significantly lower for patients with CUP compared to those with metastatic CKP: CUP median survival 14.0 months (95% C.I. 11.9 - 15.8, n = 685) vs. metastatic CKP median survival 23.1 months (95% C.I. 21.8 - 24.2, n = 7,797). This is expected as CUPs are an advanced metastatic cancer with limited treatment options [33]. The absolute difference in median survival was significant across all predicted CUP - metastatic CKP pairs with the exception of Pancreatic Adenocarcinoma (CUP PAAD median survival 8.61 months 95% C.I. 5.09 - 10.8 vs. metastatic CKP PAAD median survival 6.73 months 95% C.I. 5.98 - 8.02), known to be a particularly deadly cancer type.

### Shared prognostic somatic variants in CUP-metastatic CKP pairs

We aimed to identify prognostic somatic variants shared between OncoNPC CUP subtypes and their corresponding metastatic CKP cancers. Three out of 14 tested CUP-metastatic CKP pairs (NSCLC, PAAD, and COADREAD) exhibited shared prognostic somatic variants significantly associated with overall survival with nominal p-value cut-off at 0.05 (Fig. 4c and 4d). In patients with known or classified NSCLC, three somatic mutations were associated with poor survival in both groups: SMARCA4 (CUP: H.R. 1.86, 95% C.I. 1.19 - 2.89, p-value 6.23 × 10^−3^, CKP mets: H.R. 1.73, 95% C.I. 1.44 - 2.09, p-value 9.30 × 10^−9^), STK11 (CUP: H.R. 1.76, 95% C.I. 1.14 - 2.71, p-value 1.05 10^2^, CKP mets: H.R. 1.43, 95% C.I. 1.22 - 1.68, p-value 1.00 × 10^−5^), and KEAP1 (CUP: H.R. 1.83, 95% C.I. 1.18 - 2.85, p-value 6.82 × 10^−3^, CKP mets: H.R. 1.40, 95% C.I. 1.18 - 1.66, p-value 1.27 × 10^−4^). These associations of somatic mutations in SMARCA4, STK11, and KEAP1 genes with overall survival are well established for NSCLC [34–36]. Interestingly, a CNA event in NKX2-1 was associated with improved survival in the patients from the NSCLC pair (CUP: H.R. 0.542, 95% C.I. 0.326 - 0.901, p-value 1.83 10^2^, CKP mets: H.R. 0.770, 95% C.I. 0.662 - 0.894, p-value 6.28 × 10^−4^), consistent with prior meta-analyses [37]. In patients with known or classified COADREAD tumors, SBS10b mutation signature, linked to polymerase epsilon exonuclease domain mutations [20], was associated with longer overall survival (CUP: H.R. 0.371, 95% C.I. 0.148 - 0.928, p-value 3.41 × 10^−2^, CKP mets: H.R. 0.495, 95% C.I. 0.255 - 0.958, p-value 3.68 × 10^−2^). Finally, in patients with known or classified PAAD tumors, the SBS29 mutation signature (commonly found in tumor samples from individuals with a tobacco chewing habit [20]) was associated with poor survival in CUPs but nominally protective in metastatic CKPs (CUP: H.R. 2.66, 95% C.I. 1.02 - 6.93, p-value 4.46 × 10^−2^, CKP mets: H.R. 0.657, 95% C.I. 0.438 - 0.986, p-value 4.28 × 10^−2^). Although these somatic associations remain to be validated in independent cohorts, by categorizing patients with CUP based on their OncoNPC predictions, we were able to identify prognostic somatic variants, consistent with recent research findings.

### Identifying actionable somatic variants in CUP tumors based on OncoNPC predictions

We investigated if OncoNPC classifications could identify genetically driven, site-specific treatment options that are typically available for cancers with known primaries. We utilized OncoKB [38] as a knowledge base and considered three different categories of actionable somatic variants: oncogenic mutation, amplification, and fusion (see Methods). OncoNPC cancer type predictions enabled identification of actionable somatic variants across CUP tumor samples (total 22.8% of the eligible CUP tumor samples; see Fig. 5a and Fig. 5b). The majority of actionable somatic variants for patients with CUP were oncogenic mutations (183 counts; 87.1%), followed by amplifications (22 counts; 9.52%) and fusions (7 counts; 3.33%) as shown in Fig. 5a. The four most frequent oncogenic mutations were in PIK3CA, KRAS, ALK, and ERBB2 genes, occurring in CUP tumor samples classified as BRCA (PIK3CA and ERBB2 genes) and NSCLC (KRAS, ALK, and ERBB2 genes). Overall, among the eligible CUPs whose prediction confidences are greater than 0.5 (N = 794; see Supplementary Fig. S5 for more details on the exclusion criteria), OncoNPC predictions identified actionable somatic variants for 11.5% of the CUP tumor samples for Level 1 therapeutic level (FDAapproved drugs), 3.63% for Level 2 (Standard care), 6.64% for Level 3 (Clinical evidence), and 1.00% for Level 4 (Biological evidence), summing up to the total 22.8% of the eligible CUP tumor samples (Fig. 5b).

**Figure 5.**
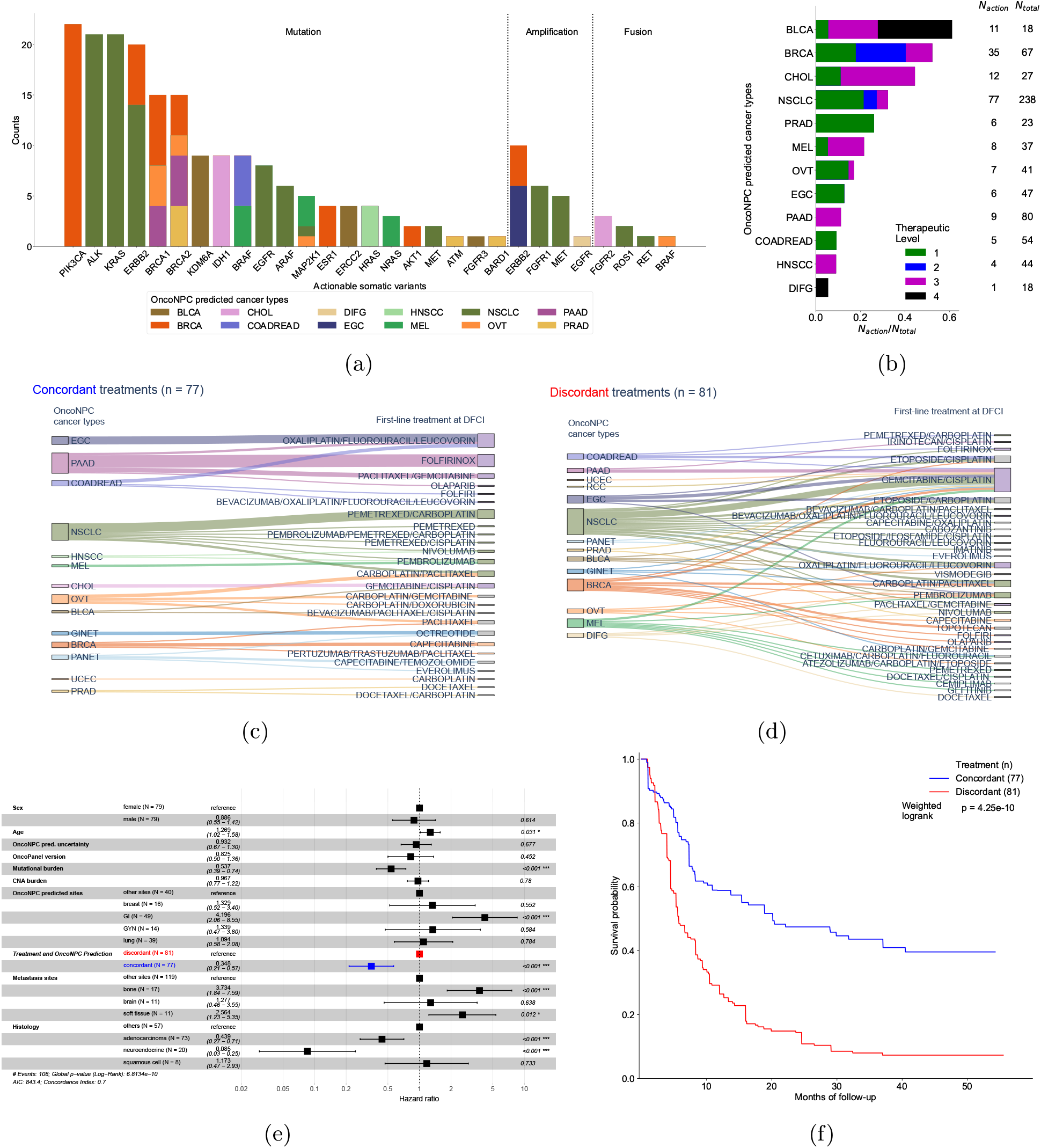
Potential for clinical decision support among OncoNPC classified CUPs. **(a)** The number of CUP tumor samples with actionable targets, based on OncoKB [38], across actionable somatic variants (mutations, amplifications, and fusions). Each bar corresponds to an actionable target, color-coded by the number of each OncoNPC classified CUP carrier. Note that each tumor sample may contain more than one actionable somatic variant. **(b)** Proportions of CUP tumor samples with actionable somatic variants (*N*_*action*_) to the total number of patients (*N*_*total*_) across OncoNPC predicted cancer types. Proportions for 4 different therapeutic levels based on OncoKB [38], are shown in each bar: Level 1 - FDA-approved drugs, Level 2 – standard of care drugs, Level 3 - drugs supported by clinical evidence, and Level 4 - drugs supported by biological evidence. **(c), (d)** Treatment diagrams for a group of patients with CUP who received treatments that were concordant with the OncoNPC classification **(c)** and the remaining CUP patients who received discordant treatments **(d)**. OncoNPC classification is shown on the left and treatment groups are shown on the right, with each patient connected from left to right. **(e)** Forest plot of a multivariable Cox Proportional Hazards Regression on patients in the CUP cohort with first-line palliative treatment records at DFCI (n = 159; see Appendix Fig. S5 for the exclusion criteria). Treatment concordance (colored in blue), encoded as 1 when the first treatment a patient received at DFCI is *concordant* with their corresponding OncoNPC prediction and 0 otherwise, was significantly associated with mortality of patients in the cohort (H.R. 0.321, 95% C.I. 0.165 - 0.620, p-value: < 0.001). **(f)** Estimated survival curves for patients with CUP in the concordant treatment group (shown in blue) and discordant treatment group (shown in red), respectively. To estimate the survival function for each group, we utilized Inverse Probability of Treatment Weighted (IPTW) Kaplan-Meier estimator while adjusting for left truncation until time of sequencing (see Methods). Statistical significance of the survival difference between the two groups was estimated by a weighted log-rank test [68].

### Survival benefit of treatment concordance with OncoNPC predictions

We performed retrospective survival analysis to investigate whether patients with CUP achieved clinical benefit when treated in concordance with their OncoNPC classifications. We restricted to a cohort of 158 patients with CUP, received first treatment at DFCI with a palliative intent (see the exclusion criteria in Supplementary Fig. S5). Each case was then manually chart reviewed by a certified oncologist to determine whether the treatment administered was concordant with the OncoNPC prediction per National Comprehensive Cancer Network (NCCN) guidelines or standard of care (see Methods, Fig. 5c, and Fig. 5d). Strikingly, patients with CUP who received first palliative treatments concordant with their OncoNPC predicted cancer types exhibited significantly better survival than those who received discordant treatments as shown in Fig. 5e and 5f (*multivariable Cox regression*: H.R. 0.348, 95% C.I. 0.210 - 0.570, p-value 2.32 × 10^−5^, Proportional Hazard assumption test [39]: Chi-squared test with 17 degrees of freedom p-value 0.156, *IPTW Kaplan-Meier estimator* : weighted log-rank test p-value 4.25 × 10^−10^). Finally, after stratifying by OncoNPC predicted cancers and repeating the IPTW Kaplan-Meier analysis, we found that the treatment concordant group had improved survival across cancer cohorts (breast, GI, and others), with the exception of the lung cancer cohort (Supplementary Fig. S7).

We note that as this was not a randomized analysis, a potential concern may be systematic differences between the concordant and discordant groups leading to a significant prognostic but not predictive difference [40]. For example, treatment discordant patients may have systematically more advanced/de-differentiated tumors and thus exhibit poorer survival regardless of their treatment regimen. (see Table 2 for comparison of the two groups across the measured covariates). To minimize biases from potential confounders and move towards a predictive estimate of treatment concordance on patient survival, we adopted two estimation strategies: multivariable Cox regression [41] (i.e., covariate adjustment) and Inverse Probability of Treatment Weighted (IPTW) Kaplan-Meier estimator [42] (see Methods), which have recently been employed to emulate estimates from randomized trials [43, 44]. In both multivariable Cox regression and IPTW Kaplan-Meier estimator strategies, patients treated like their OncoNPC predicted cancer types (i.e. those in the concordant treatment group) consistently showed significantly better survival compared to those in the discordant treatment group. The multivariable Cox regression (Fig. 5e) additionally identified significant hazardous effects of age, gastrointestinal (GI) cancer types predicted by OncoNPC, and bone metastasis (H.R. 1.27, 95% C.I. 1.02 – 1.58, p-value 3.10 × 10^− 2^, H.R. 4.20, 95% C.I. 2.06 – 8.55, p-value 7.78 × 10^− 5^, and H.R. 3.73, 95% C.I. 1.84 – 7.59, p-value 2.71 × 10^−4^, respectively), and significantly protective effects of tumor mutational burden (TMB), as well as adenocarcinoma and neuroendocrine tumor group determined by the histopathology results (H.R. 0.537, 95% C.I. 0.388 - 0.742, p-value 1.64 × 10^−4^, H.R. 0.439, 95% C.I. 0.272 - 0.710, p-value 7.85 10^4^ and H.R. 0.0854, 95% C.I. 0.0298 - 0.245, p-value 4.79 × 10^−6^, respectively). In the IPTW Kaplan-Meier analysis, we found that treatment concordance with the OncoNPC prediction was associated with Gastrointestinal (GI) cancer types (coefficient 1.916, 95% C.I. 0.627 - 3.205, p-value 3.57 × 10^−3^), whereas male sex and OncoNPC prediction uncertainty (i.e., entropy of predicted probability distribution over the considered cancer types) were inversely associated with receiving concordant treatment (coefficient -1.259, 95% C.I. -2.283 - -0.234, p-value 1.61 × 10^−2^, and coefficient -1.693, 95% C.I. -2.458 - -0.927, p-value 1.46 × 10^−5^) (see Supplementary Fig. S8). These associations with treatment concordance are consistent with likely GI CUPs being more clinically identifiable and low OncoNPC confidence CUPs being less clinically identifiable. We note, however, that the IPTW approach specifically adjusts for these systematic differences when estimating the effect of treatment concordance on survival.

**Table 2.** Demographic details of patients wit CUP in the concordant and discordant treatment groups.

|  | Concordant treatment group<br>(n = 77) | Discordant treatment group<br>(n = 81) |
| --- | --- | --- |
| Sex; male-female ratio | 0.442-0.558 | 0.556-0.444 |
| Age at sequencing (95% C.I.) | 64 (61.6 - 66.4) | 62 (59.4 - 64.6) |
| Prediction uncertainty<br>(in entropy; 95% C.I.) | 0.550 (0.426 - 0.675) | 0.988 (0.850 - 1.127) |
| **OncoPanel version (proportion in %)** |  |  |
| v1 | 1 (1.30%) | 1 (1.24%) |
| v2 | 9 (11.7%) | 15 (18.5%) |
| v3 | 67 (87.0%) | 65 (80.2%) |
| Mutational burden (95% C.I.) | 0.027 (0.021 - 0.033) | 0.033 (0.027 - 0.040) |
| CNA burden (95% C.I.) | 0.201 (0.166 - 0.236) | 0.186 (0.155 - 0.217) |
| **Predicted primary cancer groups (proportion in %)** |  |  |
| Lung | 15 (19.5%) | 24 (29.6%) |
| Breast | 5 (6.50%) | 11 (13.6%) |
| GI | 33 (42.9%) | 16 (19.8%) |
| Gyn | 9 (11.7%) | 5 (6.17%) |
| Others | 15 (19.5%) | 25 (31.0%) |
| **Metastatic sites (proportion in %)** |  |  |
| Brain | 4 (5.20%) | 8 (9.88%) |
| Bone | 7 (9.10%) | 10 (12.3%) |
| Soft tissue | 6 (7.79%) | 5 (6.17%) |
| Others | 60 (77.9%) | 58 (71.6%) |
| **Histology (proportion in %)** |  |  |
| Adenocarcinoma | 41 (53.2%) | 32 (39.5%) |
| Neuroendocrine | 9 (11.7%) | 11 (13.6%) |
| Squamous cell | 2 (2.60%) | 6 (7.41%) |
| Others | 25 (32.5%) | 32 (39.5%) |
| Treatment start date (95% C.I.) | 2018-4-30 (2017-12-24 - 2018-9-3) | 2018-3-1 (2017-10-28 - 2018-7-3) |

## Discussion

Our work provides unique insights into the genetic and prognostic landscapes of CUP tumor samples by utilizing routinely collected EHR and multicenter NGS tumor panel sequencing data. We have developed OncoNPC, a machine learning model for molecular classification of tumor samples based on the NGS panel data. When evaluated with the held-out multicenter test data, OncoNPC provided robust and interpretable predictions. Applying OncoNPC to CUP tumor samples, we demonstrated that the OncoNPC CUP subtypes showed significantly higher germline PRS risk for their predicted cancer. To our knowledge, this is the first evidence of germline genetic correlation between CUPs and corresponding known primaries, and lends orthogonal support to the molecular classification of CUPs into subtypes. We demonstrated clinical utility of the OncoNPC CUP subtypes by showing significant survival differences across subtypes, and, within subtypes, potentially actionable somatic alterations in 11.5% (Level 1 therapeutic level) and 22.8% (all levels) of tumors. Finally, in a retrospective analysis, we showed that patients with CUP, that had been treated in a consistent manner with their OncoNPC classification, achieved significantly longer survival than those treated in an inconsistent manner (multivariable Cox regression: H.R. 0.348, 95% C.I. 0.210 - 0.570, p-value 2.32 × 10^−5^). Our findings suggest that CUP tumors share a genetic and prognostic architecture with known cancer types, and may benefit from molecular classification with OncoNPC for prognosis as well as treatment decision-making.

The question of whether CUP tumors consist of heterogeneous latent primaries or are a unique cancer type in and of themselves has been actively investigated [18, 45, 46]. Prior studies have demonstrated accurate classification of known tumors using Whole-Genome Sequencing [12], NGS panels [10], RNA-seq [11], methylation [8], and other platforms [47, 48]. However, these algorithms typically applied classification to metastatic tumors of known types and did not investigate the clinical implications for CUPs at large scale. Moran et al., [8] observed a nominally significant difference in survival between patients with CUP who received site-specific treatments concordant with their molecular primary site predictions and those who received empiric treatments. While promising, it remains unknown whether this difference is due to accurate classification for the site-specific group or systematically worse outcomes for the empirically treated group, which is typically a more challenging patient population [49]. To explicitly distinguish these scenarios, our analysis instead restricted to a CUP cohort wherein all patients received site-specific treatments as the first palliative-intent therapy and estimated a significant survival benefit of concordant treatment vs. discordant treatment (excluding the empirically treated group). Our findings were obtained after adjusting for left-truncation for sequencing time and measured potential confounders through covariate adjustment as well as propensity score weighting, which have been recently employed to mimic clinical trials in Real World data [43, 44]. Although we cannot rule out potential biases from unmeasured confounders, our cohort includes more heterogeneous populations compared to recruited cohorts in randomized controlled trials (RCT), and the proposed intervention (concordant treatment vs. discordant treatment) is challenging to ethically evaluate through RCTs, necessitating the use of retrospective causal inference.

Our study has several limitations. Firstly, although we utilized multicenter NGS tumor panel sequencing data to train OncoNPC model for cancer type prediction, we utilized retrospective EHR data from a single institution for the downstream clinical analyses. As a result, these analyses may be susceptible to systematic ascertainment patterns or biases specific to a tertiary academic cancer center. Replication of our clinical findings in other institutions is thus necessary to generalize our results. Secondly, we considered only the 22 most common cancer types in the cohort as classification labels (68.1 % of all tumor samples at DFCI, and 69.9 % across all three centers). As a result, if a CUP tumor sample harbors a distinct yet not modeled primary cancer type, then the tumor sample will likely have high uncertainty in the prediction (see Supplementary Fig. S1b). Nevertheless, prior work has shown that the majority of resolvable primary sites of CUP tumor samples were from common cancers (e.g., lung, pancreas, and GI) [18], consistent with our findings. As more diverse tumor samples are collected across multiple institutions, our model can be augmented to robustly predict rare cancer types as well. Thirdly, our classifier and analyses relied on data from panel sequencing assays targeting 300-500 genes, which are inherently only sensitive to coding mutations and deep copy number alterations in the targeted genes. Other features captured by whole-genome sequencing or molecular assays may thus achieve better classification performance. Our focus in this work was on assays that are in routine clinical use as those are linked to Real World clinical data and offer the most immediate translational potential.

Our findings strongly suggest that routinely collected targeted tumor panel sequencing data have clinical utility in assisting diagnostic work-up and prognosis, and may additionally inform treatment decisions. To date, clinical sequencing is primarily used for identification of known biomarkers and corresponding clinical trial enrollment [50–53], and our findings additionally support use of panel sequencing for diagnosis. Conventional IHC-based pathology reviews are often unable to identify a primary diagnosis for advanced metastatic tumor samples [3, 6], particularly in community clinics where resources are limited. And in many cases, patients do not receive the complete diagnostic work-up that is recommended for CUPs [54]. As a result, oncologists resort to empiric treatment regimens to treat many patients with CUP [18] even when targeted therapies would otherwise be the standard of care for a corresponding known primary. In future work, we envision a multimodal framework that incorporates molecular sequencing together with patient pathology images [48], physiological data, and clinical notes to directly predict optimal treatment regiments rather than just cancer types. Our work thus paves a way for incorporating routine panel sequencing data into clinical decision support tools for clinically challenging cases.

## Methods

### Patients and tumor samples

We used the next generation sequencing (NGS) targeted panel sequencing data collected at three institutions in routine clinical care as part of the AACR project GENIE [1]: Dana-Farber Cancer Institute (DFCI, n=18,816), Memorial Sloan Kettering Cancer (MSK, n=16,294) center, and Vanderbilt-Ingram Cancer Center (VICC, n=1,335). The collected tumor samples represented 22 different cancer types and included 971 total samples from cancer of unknown primary (CUP). National Death Index (NDI) and clinical death and last clinical appointment records were available for 20,281 DFCI patients (n = 16,376 for CKP and n = 838 for CUP). Demographic details of the patients and tumor samples can be found in Table 1.

The cancer centers, DFCI, MSK, and VICC, were chosen because of similar genomic data characterization of their sequence panels in terms of coverage and alteration types [1]. DFCI samples were sequenced using a custom, hybridization-based panel called OncoPanel which targeted exons of 275-447 genes across three panel versions [1, 52]. MSK samples were sequenced using a custom panel called MSK-IMPACT which targeted 341-468 genes across 3 panel versions [1, 51]. VICC samples were sequenced using custom panels called VICC-01-T5A and VICC-01-T7, which targeted 322 and 429 genes, respectively [1]. All panels were capable of detecting single nucleotide variants (SNVs), small indels, copy number alterations, and structural variants [1].

The DFCI CUP cohort consisted of 971 sequenced tumor samples (from 962 patients) with a cancer diagnosis of CUP and the following detailed cancer type: Adenocarcinoma, Not Otherwise Specified (NOS) (n = 345), Cancer of Unknown Primary, NOS (n = 194), Squamous Cell Carcinoma, NOS (n = 114), Poorly Differentiated Carcinoma, NOS (n = 118), Neuroendocrine Tumor/Carcinoma, NOS (n = 170), Small Cell Carcinoma of Unknown Primary (n = 16), Undifferentiated Malignant Neoplasm (n = 12), and Mixed Cancer Types (n = 2). For downstream clinical analyses, we applied additional exclusion criteria, described in Supplementary Fig. S5.

### Developing OncoNPC cancer type classifier

We used a gradient tree boosting framework (XGBoost [55]) to develop OncoNPC for predicting cancer types from molecular features. In this framework, decision trees for the input features are sequentially added to an existing ensemble of the trees, such that the algorithm fits the new tree to the residuals from the ensembles with regularization on the tree structure. As the trees (a.k.a. weak learners) are added, the model learns optimal weights to combine their predictions and produces the improved outcome from the combined ensemble [55]. Owing to its high performance and scalability, the XGBoost method has been used across a wide range of applications in the healthcare space [56–58].

OncoNPC was trained and evaluated using tumors from 22 known cancer types split into 29,176 training samples and 7,289 test samples. Hyper-parameter selection was conducted using random search [59] with 10-fold cross validation within the training set while utilizing weighted F1 score as an evaluation metric. The optimal hyper-parameters were then selected and the model was evaluated on the held-out test set (n = 7,289). To predict primary sites of CUP tumors, the model was then re-trained on all CKP tumor samples and applied to the CUP tumors to estimate posterior probabilities across the 22 different cancer labels. For each tumor sample, a cancer type with the highest probability was chosen as the predicted primary site.

### Feature selection and OncoNPC model interpretation

The OncoNPC model was trained on somatic variant features from tumor sequencing data, as well as patient age at sequencing and sex. Other demographic/clinical features were intentionally not used so as not to bias the model toward cancer types with more available information. Somatic variant features included: mutations (i.e., single nucleotide variants (SNV) and indels), Copy Number Alteration (CNA) events, and mutational signatures [60]. For each gene, the total count of a somatic mutation (i.e., single nucleotide variants and indels) was encoded as a positive integer feature. The presence of a CNA event for each gene was encoded as a categorical variable with 5 levels: -2 (deep loss), -1 (single-copy loss), 0 (no event), 1 (low-level gain), and 2 (high-level amplification); note that CNA events data for tumor samples from MSK and VICC were encoded as -2 (deep loss), 0 (no event), and 2 (high-level amplification). Each of 60 different mutation signatures was inferred as the dot product of the weights derived from [60] and 96 single base substitutions in a trinucleotide context. The single base substitutions were computed using the deconstructSigs R library [61]. See Supplementary Table S1 for the full set of features.

To identify important features in the OncoNPC’s predictions, we used the recently proposed feature interpretation tool for tree-based models, called TreeExplainer [19] (Python package shap). TreeExplainer uses an efficient polynomial time algorithm (*O(TLD*^2^), *T* : number of trees, *L* : number of leaves, *D* : maximum depth) to approximate Shapley values which capture the impact of each feature on each individual model prediction. The Shapley value assigned to each feature is modeled as the average change in the model’s conditional expectation function over all possible feature orderings when introducing the corresponding feature into the model; it is formulated as 𝔼_*S*_ [*f*(*X*)| do (*X*_*S*_ = *x*_*S*_)], where *S* is the set of features, *X* is a random variable for the feature to perturb, and do notation [62] reflects the causal feature perturbation formulation. See [19] for more details on the algorithm and its properties.

Applying TreeExplainer on the model outcome at each fold across the 10-fold cross-fitting procedure, we obtained out-of-sample local explanations for all individual model predictions of primary cancer types. By combining local explanations of correct predictions for each cancer type, we characterized the cancer type in terms of the most important or predictive features based on their Shapley values, which provided insights into the somatic variants and clinical features most relevant to the classification of each cancer type.

### Germline PRS-based validation on CUP tumor samples

To validate the OncoNPC predictions for CUP tumor samples (which do not otherwise have a ground truth), we utilized germline Polygenic Risk Scores (PRS) which were never available to OncoNPC for training. Germline imputation from the off-target tumor sequencing data was conducted as previously described in [63]. Using weights from external GWAS data, we imputed PRS for Non-Small Cell Lung Cancer (NSCLC), Invasive Breast Carcinoma (BRCA), Colorectal Adenocarcinoma (COADREAD), Diffuse Glioma (DIFG), Melanoma (MEL), Ovarian Epithelial Tumor (OVT), Renal Cell Carcinoma (RCC), and Prostate Adenocarcinoma (PRAD). Pearson correlation between the PRS from off-target tumor data versus matched germline SNP array was previously shown to be higher than 0.9 without observable outliers [63].

We hypothesized that germline PRS specific to the underlying primary cancer type of a CUP tumor sample would be enriched in a manner similar to how the PRS specific to CKP tumor sample with the same primary cancer type is enriched. To that end, given the set of 8 different cancer types 𝒞 we have the imputed PRS available for, we first restricted the cohort of CUP tumor samples to those with OncoNPC predictions in 𝒞(*N*_CUP,𝒞_ = 505). Then, we obtained standardized germline PRS values for the chosen CUP tumor samples over all the cancer types in 𝒞. Finally, we defined 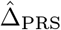 as the estimated mean difference between the PRS specific to the predicted primary cancer type 𝒞 (i.e. concordant PRS; PRS_*C*_) and average of PRSs corresponding to the rest of the cancer types (i.e. discordant PRS; PRS_*D*_, where *D* ∈ 𝒞 \ 𝒞) as follows

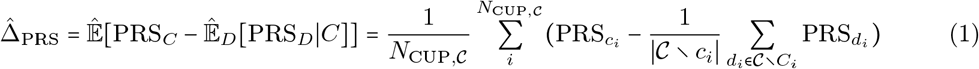

. As a true positive reference, we repeated the above procedure for the CKP tumor samples. Finally, as a true negative null, we estimated 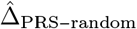, where the concordant cancer type was randomly assigned. We then repeated the random assignment 100 times to obtain estimated mean and standard errors.

### Survival function estimation

National Death Index (NDI) and in-house clinical records were available for 20,281 DFCI patients (n = 16,376 for CKP and n = 838 for CUP). A patient’s lost to follow-up date was determined at either the last NDI update date (12/31/2020) or their corresponding last contact date from the in-house records, whichever date is later. A patient’s death date was determined from the in-house records, or the NDI data if the patient was lost to follow-up.

#### CUP-metastatic CKP survival comparison

We estimated median survival times of patients across CUP - metastatic CKP pairs using the Kaplan-Meier estimator [64] to account for patients lost to follow-up. For the CUP cohort, we excluded patients with CUP that were lost to follow up at the time of tumor sequencing and those whose primary cancer types were predicted with low probability (see Supplementary Fig. S5). The resulting CUP cohort (n = 685), was then restricted to OncoNPCcancer types with more than 35 CUP patients. For the CKP metastatic cohort, we excluded patients lost to follow up at the tumor sequencing time in the same manner and chose patients with one of the known cancers, where either the biopsy was metastatic or the patient had an ICD-10 code indicative of secondary malignant neoplasms within a year prior to sequencing dates. A total of 521 and 5,937 patients were thus retained from the CUP cohort and metastatic CKP cohort, respectively: Non-Small Cell Lung Cancer (NSCLC; n_CUP_ = 200, n_met-CKP_ = 1,559), Pancreatic Adenocarcinoma (PAAD; n_CUP_ = 80, n_met-CKP_ = 357), Invasive Breast Carcinoma (BRCA; n_CUP_ = 67, n_met-CKP_ = 1,656), Colorectal Adenocarcinoma (COADREAD; n_CUP_ = 54, n_met-CKP_ = 1,198), Head and Neck Squamous Cell Carcinoma (HNSCC; n_CUP_ = 44, n_met-CKP_ = 216), Esophagogastric Adenocarcinoma (EGC; n_CUP_ = 40, n_met-CKP_ = 336), and Ovarian Epithelial Tumor (OVT; n_CUP_ = 36, n_met-CKP_ = 615). Note that patients with CUP, whose predicted cancer type is Gastrointestinal Neuroendocrine Tumors (GINET; n_CUP_ = 39, n_CKP_ = 118), were excluded due to the fact that the estimated survival function for the CUP cohort never reached 50 percent.

#### OncoNPC-based risk stratification among patients with CUP

To identify OncoNPC CUP subtypes with significant prognostic differences, we estimated survival functions for 7 common OncoNPC subtypes with more than 35 CUP patients: NSCLC, PAAD, BRCA, HNSCC, EGC, GINET, and Pancreatic Neuroendocrine Tumor (PANET). Patients that were lost to follow up at time of sequencing were again excluded, as were CUPs with an OncoNPC prediction probability lower than 0.5 (i.e., same criteria as the CUP - metastatic CKP survival comparison analysis). We merged subtypes with similar morphology and estimated survival functions: PAAD and EGC; GINET and PANET. To statistically test survival differences between these 5 groups, we utilized Chi-squared test with 4 degrees of freedom.

### Identifying prognostic somatic variants shared in CUP-metastatic CKP pairs

To identify prognostic somatic variants shared between CUP/metastatic-CKP pairs, we again restricted to the 7 common OncoNPC subtypes with at least 35 CUP patients: NSCLC, PAAD, BRCA, COADREAD, HNSCC, EGC, GINET, and OVT. For somatic variants, we utilized the same processed features utilized in the OncoNPC model training (see Methods: Feature selection and OncoNPC model interpretation). To ensure sufficient statistical power, we restricted to candidate somatic variants (i.e., mutated genes and CNA genes) present in at least 15 samples in a given OncoNPC subtype and corresponding metastatic CKP cohort, as well as all 96 mutational signatures.

After selecting the cancer types to consider in the CUP-metastatic CKP pairs and candidate somatic variants for each pair, we iteratively tested each feature for association with survival in each OncoNPC subtype and in each corresponding metastatic CKP cohort. A multivariable Cox Proportional Hazard regression [41] model was used with time-to-death from sequencing as the outcome. To adjust for baseline effects, we included age at sequencing, sex, tumor sequencing panel version, mutational burden (i.e., sum of total somatic mutations in each tumor sample), and CNA burden (i.e., sum of total CNA events in each tumor sample) as covariates. Finally, to identify shared prognostic somatic variants for each CUP-metastatic CKP pair, we retained somatic variants which passed Schoenfield residuals-based proportional hazard tests (`lifelines` Python library [65]: p-value threshold: 0.05) and were nominally significant (p < 0.05) for both CUP and CKP cancer types in each pair.

### Actionable somatic variants in CUP tumors

We estimated the frequency of known, actionable somatic alterations in each OncoNPC CUP subtype using the OncoKB knowledge base [38]. OncoNPC CUP predictions with a probability greater than were retained (see Supplementary Fig. S5). We considered 3 different types for somatic variants: oncogenic mutations such as indels, missense mutations, and splice site mutations, amplifications such as high-level amplifications, and finally fusions such as gene-gene and gene-intergenic fusions as specified in OncoKB. For each actionable somatic variant, we assigned one of the four therapeutic levels: level 1 for FDA-approved drugs, level 2 for standard care drugs, level 3 for drugs supported by clinical evidence, and level 4 for drugs supported by biological evidence.

### Estimating impacts of treatment concordance on survival of patients with CUP

We estimated the impact of the concordance between treatment and OncoNPC CUP predictions on a mortality outcome in a retrospective survival analysis. We utilized the in-house patient follow-up and treatment data to identify patients with CUP who received first treatment at DFCI with a palliative intent (Supplementary Fig. S5 for the exclusion criteria). Each patient was reviewed by a trained oncologist to determine whether the OncoNPC predicted cancer type was concordant or discordant with the first line of treatment received, per National Comprehensive Cancer Network (NCCN) guidelines or standard of care, in most reasonable situations, and within the clinical context delineated in the medical record. See Supplementary Section: *Determining treatment-OncoNPC concordance* for more details, and Supplementary Table S3 for clinical information, including primary cancer diagnosis, biopsy site, and first chemotherapy plan at DFCI, of patients with CUP in the analysis.

As we were interested in the counterfactual causal impact of the OncoNPC-treatment concordance, we utilized the principles of causal inference to account for potential patient heterogeneity and confounding. Specifically, we estimated the effect of treatment concordance specified by the indicator variable, *A*, which was 1 when the first palliative treatment for a patient with CUP was concordant with the corresponding OncoNPC prediction and 0 otherwise. Our analyses make the following identifiability assumptions:

- Conditional ignorability : 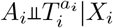, where *A*_*i*_ ∈ 0, 1. It means that given patient *i*’s a set of covariates *X*_*i*_, the patient’s treatment concordance *A*_*i*_ is as good as random.
- Consistency : 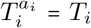, which means that a counterfactual outcome 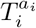*i* for patient *i* is the observed outcome for the patient with a treatment concordance *a*_*i*_.
- Overlap : *P*(0 < *p* (*X*_*i*_) < 1) = 1 where *p* (*X*_*i*_) = *P*(*A*_*i*_ = 1| *X*_*i*_), which means all patients have a strictly positive probability for receiving concordant treatment (*A*_*i*_ = 1).

In addition to the above identifiability assumptions, we made independent censoring (i.e. *C*_*i*_ ⊧*T*_*i*_|*X*_*i*_) and independent entry assumption given the covariates (i.e. *E*_*i*_ *T*_*i*_|*X*_*i*_).

We adopted two different estimation strategies to obtain the impact of treatment concordance: semi-parametric Cox Proportional Hazard estimator adjusted with a set of measured confounders *X* [41] and non-parametric Kaplan Meier estimator adjusted with Inverse Probability Treatment Weighting (IPTW). We formulated an IPTW, *w*_*i*_ for each sample as 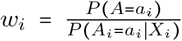 [42] and estimated *P A* non-parametrically and *P A X* using a logistic regression model (R `glm` package [66]) in a 10-fold cross-fitting. A set of measured confounders (i.e., *X*_*i*_) included patients’ sex, age, OncoNPC prediction uncertainty (in entropy of posterior distribution over 22 cancer types), sequencing panel (i.e., OncoPanel) version, mutational burden, CNA burden, subsets of OncoNPC predicted cancer types and metastasis sites, and finally pathological histology (e.g., adenocarcinoma tumor or neuroendocrine tumor). Since patients with CUP who met the treatment criteria (i.e., follow-up start time) but did not receive clinical panel sequencing (i.e., entry time) could not be included in the analysis, we adjusted for the left truncation by defining the risk set ℛ(*t*) at time *t*, which corresponds to the set of patients followed up in the analysis up to time *t* as follows

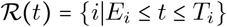

, where *E*_*i*_ is the entry time of patient *i*. With the independent entry assumption as stated before, we obtained survival function from Kaplan-Meier estimator as follows

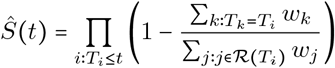

. In this formulation, each individual is weighted by the corresponding IPTW, *w*_*i*_, and we obtained two different survival functions for the treatment concordant and discordant groups. The adjusted Kaplan-Meier estimator provides a consistent estimate of impact of the treatment concordance under the assumptions stated above [42]. Once we obtained the survival estimates for the two groups, we used a weighted log-rank test [67] to test for a significant difference in survival.

In the Cox proportional hazard regression framework, we estimated the hazard function of patient *i* as follows: *λ* (*t* | *A*_*i*_, *X*_*i*_)) *= λ*_0_ (*t*) exp (*αA*_*i*_ *+ β*^*T*^ *X*_*i*_), where *α, A*_*i*_ ∈ ℝ and *β, X*_*i*_ ∈ ℝ^*m*^ (*m* is the number of measured confounders). Under the above identifiability assumptions and validity of the estimation model, *e*^*α*^ is the hazard ratio capturing the causal effect of the treatment concordance *A*. Finally, under the assumption of no ties between event times across the patients, the parameters *α* and *β* are estimated by maximizing the following partial likelihood

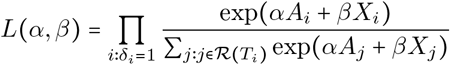

[41].

## Supporting information

All supplementary materials

## Data Availability

All data produced in the present study are available upon reasonable request to the authors.

https://github.com/itmoon7/onconpc

## Acknowledgments

The participation of patients and the efforts of an institutional data collection system made this study possible, and we are grateful for their contributions. We would also like to express our appreciation to the DFCI Oncology Data Retrieval System (OncDRS) and AACR Project GENIE team for their role in aggregating, managing, and delivering the data used in this project.

## Supplementary Note

### Determining treatment-OncoNPC concordance

Concordance of OncoNPC predicted cancer type with a first palliative treatment assignments at DFCI was classified in one of five categories: 1) “TRUE”: the OncoNPC cancer type matched the clinically proven/suspected tumor type and the predicted treatment matched the treatment received, which was dictated by NCCN guidelines and/or standard of care, within the clinical context provided by the medical record; 2) “FALSE”: the OncoNPC cancer type did not match the clinically proven/suspected cancer type and the predicted treatment was not appropriate per NCCN guidelines or standard of care, in most reasonable situations, and within the context of the medical record; 3) “SOFT FALSE”: the OncoNPC cancer type did not match the clinically proven/suspected cancer type, but the treatment received was not chosen based on NCCN guidelines or standard of care, owing to the unique clinical context provided by the medical record, 4) “EMPIRIC”: treatment received was empiric treatment for cancer of unknown primary (e.g., carboplatin/taxol or gemcitabine/cisplatin) with the corresponding clinical rationale; in cases where patients received these regimens but not with the clinical intent of empiric CUP treatment (i.e., as regimens intended for treating other tumor types), the predicted treatment was not labeled as “EMPIRIC” and the case was instead evaluated in context of the proven/suspected tumor type. In our analysis, we considered the TRUE group as the concordant group, and FALSE and SOFT FALSE groups as the discordant group. We did not include the EMPIRIC group, which is typically a more challenging patient population with systematically worse outcomes [49].

### Code Availability

Please see https://github.com/itmoon7/onconpc for the pre-processing script, the trained OncoNPC model, and other reference materials.

## Figure and Table Legends

**Supplementary Figure S1. OncoNPC prediction performances and confidences (i.e**., *p*_**max**_**) across centers. (a)** Center-specific OncoNPC performance (in weighted F1) on the test CKP tumor samples (n = 7,289). The figure is a decomposed version of Fig. 2c by cancer center (DFCI: ◯, MSK: ◻, VICC: ◇). The performance was evaluated at 4 different prediction confidences (i.e., minimum *p*_*max*_ thresholds). Each dot size is scaled by the proportion of tumor samples retained. See Table S2 for the center-specific number of test CKP tumor samples across cancer types. **(b), (c)** Box plots of prediction confidences (*p*_max_) across **(b)** DFCI CUP tumors, MSK CUP tumors, all DFCI CKP tumors, DFCI held-out CKP tumors, and DFCI excluded CKP tumors, and **(c)** DFCI held-out CKP tumors, MSK held-out CKP tumors, and VICC held-out CKP tumors. Medians and lower and upper quartiles are shown on the figures along with corresponding number of tumor samples as well as means and 95% confidence intervals.

**Supplementary Figure S2. Interpreting OncoNPC predictions**. Top 15 most important features based on mean absolute SHAP values (i.e., 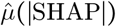 [19]) for cancer types with at least 20 CUP tumors samples were classified into.

**Supplementary Figure S3. SHAP summary plot** [19] for cancer types with at least 20 CUP tumors samples were classified into. SAHP values (i.e., impact on OncoNPC predictions) are shown on the x-axis, while features values are shown with a color map (from purple to yellow). In each plot, CUP and CKP tumor samples were combined into one cohort for the corresponding cancer.

**Supplementary Figure S4. Applying OncoNPC to MSK CUP tumor samples. (a)** Empirical distributions of prediction probabilities for correctly predicted, held-out CKP tumor samples (n = 3,429) and MSK CUP tumor samples (n = 496) across CKP cancer types (blue) and their corresponding OncoNPC predicted cancer types for CUP tumors (green). Only OncoNPC classifications with at least 20 CUP tumor samples are shown. **(b)** Proportion of each CKP cancer type and the corresponding OncoNPC predicted CUP cancer type. All training CKP tumor samples (n = 36,445) and all MSK CUP tumor samples (n = 581) are shown. For both **(a)** and **(b)**, the cancer types (x-axis) are ordered by the number of CKP tumor samples in each cancer type.

**Supplementary Figure S5**. Exclusion criteria for downstream clinical analyses.

**Supplementary Figure S6**. Germline Polygenic Risk Score (PRS) enrichment of CKP tumor samples and CUP tumor samples across 8 different cancer types: **(a)** Colorectal Adenocarcinoma (COADREAD), **(b)** Diffuse Glioma (DIFG), **(c)** Invasive Breast Carcinoma (BRCA), **(d)** Melanoma (MEL), **(e)** Non-Small Cell Lung Cancer (NSCLC), **(f)** Ovarian Epithelial Tumor (OVT), **(g)** Prostate Adenocarcinoma (PRAD), and **(h)** Renal Cell Carcinoma (RCC). The magnitude of the enrichment is quantified by 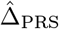: the mean difference between the concordant (i.e. OncoNPC matching) cancer type PRS and mean of PRSs of discordant cancer types (see Methods). 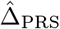 is shown for CKPs in blue (for reference) and CUPs in green.

**Supplementary Figure S7**. Estimated survival curves for patients with CUP, broken down by OncoNPC predicted cancer types: **(a)** BRCA, **(b)** Gastrointestinal (GI) group (CHOL, COAD-READ, EGC, and PAAD), **(c)** Lung (NSCLC and PLMESO), and **(d)** other OncoNPC cancer types (BLCA, DIFG, GINET, HNSCC, MEL, OVT, PANET, PRAD, RCC, and UCEC). In each figure, the concordant treatment group and discordant treatment group are shown in blue and red, respectively. To estimate the survival function for each group, we utilized Inverse Probability of Treatment Weighted (IPTW) Kaplan-Meier estimator while adjusting for left truncation until time of sequencing (see Methods). Statistical significance of the survival difference between the two groups was estimated by a weighted log-rank test [68].

**Supplementary Figure S8. Summary of coefficients for estimating treatment-OncoNPC concordance**. Formally, we estimated out-of-sample *P* (*A* | *X*), where *A* corresponds to the treatment-OncoNPC concordance, using a logistic regression model in a 10-fold cross-fitting. The coefficients were obtained from the first fold. See Methods: Estimating impacts of treatment concordance on survival of patients with CUP for more details.

**Supplementary Table S1**. A full set of 861 somatic input features for OncoNPC, abstracted from the next generation NGS targeted panel sequencing data. The features belong to three different categories (shown as columns of the table): somatic mutations (i.e., single nucleotide variants and indels: 316 features), Copy Number Alterations (CNA: 491 features), and mutational signatures (54 features). Note that we included patients’ sex and age in addition to the somatic features. See Methods for more details on how the features were encoded.

**Supplementary Table S2**. Center-specific number of held-out CKP tumor samples across cancer types and prediction confidence (i.e., *p*_max_) thresholds.

**Supplementary Table S3**. Clinical information of patient with CUP in the treatment concordance analysis (n = 158).

## References

[1] A. P. G. Consortium et al., “Aacr project genie: Powering precision medicine through an international consortium,” Cancer discovery, vol. 7, no. 8, pp. 818–831, 2017.

[2] N. Pavlidis, H. Khaled, and R. Gaafar, “A mini review on cancer of unknown primary site: A clinical puzzle for the oncologists,” Journal of advanced research, vol. 6, no. 3, pp. 375–382, 2015.

[3] G. R. Varadhachary and M. N. Raber, “Cancer of unknown primary site,” New England Journal of Medicine, vol. 371, no. 8, pp. 757–765, 2014.

[4] D. M. Hyman et al., “Vemurafenib in multiple nonmelanoma cancers with braf v600 mutations,” New England Journal of Medicine, vol. 373, no. 8, pp. 726–736, 2015.

[5] J. D. Hainsworth and F. A. Greco, “Cancer of unknown primary site: New treatment paradigms in the era of precision medicine,” American Society of Clinical Oncology Educational Book, vol. 38, pp. 20–25, 2018.

[6] G. G. Anderson and L. M. Weiss, “Determining tissue of origin for metastatic cancers: Meta-analysis and literature review of immunohistochemistry performance,” Applied Immunohistochemistry & Molecular Morphology, vol. 18, no. 1, pp. 3–8, 2010.

[7] K. A. Oien and J. L. Dennis, “Diagnostic work-up of carcinoma of unknown primary: from immunohistochemistry to molecular profiling,” Ann Oncol, vol. 23 Suppl 10, pp. x271–277, Sep. 2012.

[8] S. Moran et al., “Epigenetic profiling to classify cancer of unknown primary: A multicentre, retrospective analysis,” The Lancet Oncology, vol. 17, no. 10, pp. 1386–1395, 2016.

[9] W. Jiao et al., “A deep learning system can accurately classify primary and metastatic cancers based on patterns of passenger mutations,” bioRxiv, p. 214 494, 2019.

[10] A. Penson et al., “Development of genome-derived tumor type prediction to inform clinical cancer care,” JAMA oncology, vol. 6, no. 1, pp. 84–91, 2020.

[11] B. He et al., “A neural network framework for predicting the tissue-of-origin of 15 common cancer types based on rna-seq data,” Frontiers in Bioengineering and Biotechnology, vol. 8, p. 737, 2020.

[12] L. Nguyen, A. Van Hoeck, and E. Cuppen, “Machine learning-based tissue of origin classification for cancer of unknown primary diagnostics using genome-wide mutation features,” Nature communications, vol. 13, 2022.

[13] J. D. Hainsworth et al., “Molecular gene expression profiling to predict the tissue of origin and direct site-specific therapy in patients with carcinoma of unknown primary site: A prospective trial of the sarah cannon research institute,” Journal of Clinical Oncology, vol. 31, no. 2, pp. 217–223, 2013.

[14] H. Yoon et al., “Gene expression profiling identifies responsive patients with cancer of unknown primary treated with carboplatin, paclitaxel, and everolimus: Ncctg n0871 (alliance),” Annals of Oncology, vol. 27, no. 2, pp. 339–344, 2016.

[15] H. Hayashi et al., “Site-specific and targeted therapy based on molecular profiling by next-generation sequencing for cancer of unknown primary site: A nonrandomized phase 2 clinical trial,” JAMA oncology, vol. 6, no. 12, pp. 1931–1938, 2020.

[16] H. Hayashi et al., “Randomized phase ii trial comparing site-specific treatment based on gene expression profiling with carboplatin and paclitaxel for patients with cancer of unknown primary site,” Journal of Clinical Oncology, vol. 37, no. 7, pp. 570–579, 2019.

[17] A.-M. Conway, C. Mitchell, and N. Cook, “Challenge of the unknown: How can we improve clinical outcomes in cancer of unknown primary?” Journal of clinical oncology: official journal of the American Society of Clinical Oncology, vol. 37, no. 23, pp. 2089–2090, 2019.

[18] T. Bochtler and A. Krämer, “Does cancer of unknown primary (cup) truly exist as a distinct cancer entity?” Frontiers in oncology, vol. 9, p. 402, 2019.

[19] S. M. Lundberg et al., “From local explanations to global understanding with explainable ai for trees,” Nature machine intelligence, vol. 2, no. 1, pp. 56–67, 2020.

[20] J. G. Tate et al., “Cosmic: The catalogue of somatic mutations in cancer,” Nucleic acids research, vol. 47, no. D1, pp. D941–D947, 2019.

[21] G. da Cunha Santos, F. A. Shepherd, and M. S. Tsao, “Egfr mutations and lung cancer,” Annual Review of Pathology: Mechanisms of Disease, vol. 6, pp. 49–69, 2011.

[22] Y.-L. Zhang et al., “The prevalence of egfr mutation in patients with non-small cell lung cancer: A systematic review and meta-analysis,” Oncotarget, vol. 7, no. 48, p. 78 985, 2016.

[23] S. S. Hecht, “Tobacco smoke carcinogens and lung cancer,” JNCI: Journal of the National Cancer Institute, vol. 91, no. 14, pp. 1194–1210, 1999.

[24] R. Mehra et al., “Identification of gata3 as a breast cancer prognostic marker by global gene expression meta-analysis,” Cancer research, vol. 65, no. 24, pp. 11 259–11 264, 2005.

[25] S. Elsheikh et al., “Ccnd1 amplification and cyclin d1 expression in breast cancer and their relation with proteomic subgroups and patient outcome,” Breast cancer research and treatment, vol. 109, no. 2, pp. 325–335, 2008.

[26] X. Liu, M. Jakubowski, and J. L. Hunt, “Kras gene mutation in colorectal cancer is correlated with increased proliferation and spontaneous apoptosis,” American journal of clinical pathology, vol. 135, no. 2, pp. 245–252, 2011.

[27] D. Dinu et al., “Prognostic significance of kras gene mutations in colorectal cancer-preliminary study,” Journal of medicine and life, vol. 7, no. 4, p. 581, 2014.

[28] R. Hayes, J. Van Nieuwenhuize, J. Raatgever, and F. Ten Kate, “Aflatoxin exposures in the industrial setting: An epidemiological study of mortality,” Food and Chemical Toxicology, vol. 22, no. 1, pp. 39–43, 1984.

[29] M. A. Ahmed Adam, Y. M. Tabana, K. B. Musa, and D. A. Sandai, “Effects of different mycotoxins on humans, cell genome and their involvement in cancer,” Oncology Reports, vol. 37, no. 3, pp. 1321–1336, 2017.

[30] S. Marchese, A. Polo, A. Ariano, S. Velotto, S. Costantini, and L. Severino, “Aflatoxin b1 and m1: Biological properties and their involvement in cancer development,” Toxins, vol. 10, no. 6, p. 214, 2018.

[31] P. M. Westcott and M. D. To, “The genetics and biology of kras in lung cancer,” Chinese journal of cancer, vol. 32, no. 2, p. 63, 2013.

[32] H. Adderley, F. H. Blackhall, and C. R. Lindsay, “Kras-mutant non-small cell lung cancer: Converging small molecules and immune checkpoint inhibition,” EBioMedicine, vol. 41, pp. 711–716, 2019.

[33] A. M. Conway, C. Mitchell, E. Kilgour, G. Brady, C. Dive, and N. Cook, “Br J CancerMolecular characterisation and liquid biomarkers in Carcinoma of Unknown Primary (CUP): taking the ‘U’ out of ‘CUP’,” Br J Cancer, vol. 120, no. 2, pp. 141–153, Jan. 2019.

[34] A. J. Schoenfeld et al., “The genomic landscape of smarca4 alterations and associations with outcomes in patients with lung cancersmarca4 alterations in lung cancer,” Clinical Cancer Research, vol. 26, no. 21, pp. 5701–5708, 2020.

[35] S. Papillon-Cavanagh, P. Doshi, R. Dobrin, J. Szustakowski, and A. M. Walsh, “Stk11 and keap1 mutations as prognostic biomarkers in an observational real-world lung adenocarcinoma cohort,” ESMO open, vol. 5, no. 2, e000706, 2020.

[36] T. Takahashi et al., “Mutations in keap1 are a potential prognostic factor in resected non-small cell lung cancer,” Journal of surgical oncology, vol. 101, no. 6, pp. 500–506, 2010.

[37] T. Berghmans et al., “Thyroid transcription factor 1—a new prognostic factor in lung cancer: A meta-analysis,” Annals of oncology, vol. 17, no. 11, pp. 1673–1676, 2006.

[38] D. Chakravarty et al., “Oncokb: A precision oncology knowledge base,” JCO precision oncology, vol. 1, pp. 1–16, 2017.

[39] P. M. Grambsch and T. M. Therneau, “Proportional hazards tests and diagnostics based on weighted residuals,” Biometrika, vol. 81, no. 3, pp. 515–526, 1994.

[40] L. Simms, H. Barraclough, and R. Govindan, “Biostatistics primer: What a clinician ought to know—prognostic and predictive factors,” Journal of Thoracic Oncology, vol. 8, no. 6, pp. 808–813, 2013.

[41] D. R. Cox, “Regression models and life-tables,” Journal of the Royal Statistical Society: Series B (Methodological), vol. 34, no. 2, pp. 187–202, 1972.

[42] J. Xie and C. Liu, “Adjusted kaplan–meier estimator and log-rank test with inverse probability of treatment weighting for survival data,” Statistics in medicine, vol. 24, no. 20, pp. 3089–3110, 2005.

[43] R. Liu et al., “Systematic pan-cancer analysis of mutation–treatment interactions using large real-world clinicogenomics data,” Nature Medicine, vol. 28, no. 8, pp. 1656–1661, 2022.

[44] R. Liu et al., “Evaluating eligibility criteria of oncology trials using real-world data and ai,” Nature, vol. 592, no. 7855, pp. 629–633, 2021.

[45] S. Kolling et al., ““metastatic cancer of unknown primary” or “primary metastatic cancer”?” Frontiers in Oncology, vol. 9, p. 1546, 2020.

[46] T. Olivier et al., “Redefining cancer of unknown primary: Is precision medicine really shifting the paradigm?” Cancer treatment reviews, vol. 97, p. 102 204, 2021.

[47] E. Moiso et al., “Developmental deconvolution for classification of cancer origin,” medRxiv, 2021.

[48] M. Y. Lu et al., “Ai-based pathology predicts origins for cancers of unknown primary,” Nature, vol. 594, no. 7861, pp. 106–110, 2021.

[49] K. Fizazi, F. Greco, N. Pavlidis, G. Daugaard, K. Oien, and G. Pentheroudakis, “Cancers of unknown primary site: Esmo clinical practice guidelines for diagnosis, treatment and followup,” Annals of Oncology, vol. 26, pp. v133–v138, 2015.

[50] T. L. Stockley et al., “Molecular profiling of advanced solid tumors and patient outcomes with genotype-matched clinical trials: The princess margaret impact/compact trial,” Genome medicine, vol. 8, no. 1, pp. 1–12, 2016.

[51] D. T. Cheng et al., “Memorial sloan kettering-integrated mutation profiling of actionable cancer targets (msk-impact): A hybridization capture-based next-generation sequencing clinical assay for solid tumor molecular oncology,” The Journal of molecular diagnostics, vol. 17, no. 3, pp. 251–264, 2015.

[52] E. P. Garcia et al., “Validation of oncopanel: A targeted next-generation sequencing assay for the detection of somatic variants in cancer,” Archives of Pathology and Laboratory Medicine, vol. 141, no. 6, pp. 751–758, 2017.

[53] D. Sha, Z. Jin, J. Budczies, K. Kluck, A. Stenzinger, and F. A. Sinicrope, “Tumor mutational burden as a predictive biomarker in solid tumors,” Cancer discovery, vol. 10, no. 12, pp. 1808–1825, 2020.

[54] L. Mileshkin et al., “Cancer-of-unknown-primary-origin: A seer–medicare study of patterns of care and outcomes among elderly patients in clinical practice,” Cancers, vol. 14, no. 12, p. 2905, 2022.

[55] T. Chen and C. Guestrin, “Xgboost: A scalable tree boosting system,” in Proceedings of the 22nd acm sigkdd international conference on knowledge discovery and data mining, 2016, pp. 785–794.

[56] Y. Chen et al., “Physiol MeasClassification of short single-lead electrocardiograms (ECGs) for atrial fibrillation detection using piecewise linear spline and XGBoost,” Physiol Meas, vol. 39, no. 10, p. 104 006, Oct. 2018.

[57] C. M. Hatton, L. W. Paton, D. McMillan, J. Cussens, S. Gilbody, and P. A. Tiffin, “Predicting persistent depressive symptoms in older adults: A machine learning approach to personalised mental healthcare,” Journal of affective disorders, vol. 246, pp. 857–860, 2019.

[58] A. Ogunleye and Q.-G. Wang, “Xgboost model for chronic kidney disease diagnosis,” IEEE/ACM transactions on computational biology and bioinformatics, vol. 17, no. 6, pp. 2131–2140, 2019.

[59] J. Bergstra and Y. Bengio, “Random search for hyper-parameter optimization.,” Journal of machine learning research, vol. 13, no. 2, 2012.

[60] L. B. Alexandrov et al., “NatureThe repertoire of mutational signatures in human cancer,” Nature, vol. 578, no. 7793, pp. 94–101, Feb. 2020.

[61] R. Rosenthal, N. McGranahan, J. Herrero, B. S. Taylor, and C. Swanton, “Genome BiolD-econstructSigs: delineating mutational processes in single tumors distinguishes DNA repair deficiencies and patterns of carcinoma evolution,” Genome Biol, vol. 17, p. 31, Feb. 2016.

[62] D. Janzing, L. Minorics, and P. Blöbaum, “Feature relevance quantification in explainable ai: A causal problem,” in International Conference on artificial intelligence and statistics, PMLR, 2020, pp. 2907–2916.

[63] A. Gusev, S. Groha, K. Taraszka, Y. R. Semenov, and N. Zaitlen, “Constructing germline research cohorts from the discarded reads of clinical tumor sequences,” Genome medicine, vol. 13, no. 1, pp. 1–14, 2021.

[64] E. L. Kaplan and P. Meier, “Nonparametric estimation from incomplete observations,” Journal of the American statistical association, vol. 53, no. 282, pp. 457–481, 1958.

[65] C. Davidson-Pilon, “Lifelines: Survival analysis in python,” Journal of Open Source Software, vol. 4, no. 40, p. 1317, 2019.

[66] I. Marschner, M. W. Donoghoe, and M. M. W. Donoghoe, “Package ‘glm2’,” Journal, Vol, vol. 3, no. 2, pp. 12–15, 2018.

[67] A. Pezzi, M. Cavo, A. Biggeri, E. Zamagni, and O. Nanni, “Inverse probability weighting to estimate causal effect of a singular phase in a multiphase randomized clinical trial for multiple myeloma,” BMC medical research methodology, vol. 16, no. 1, pp. 1–10, 2016.

[68] S. Xu et al., “Extension of kaplan-meier methods in observational studies with time-varying treatment,” Value in Health, vol. 15, no. 1, pp. 167–174, 2012.

