## Supplementary material for "Utilizing Electronic Health Records (EHR) and Tumor Panel Sequencing to Demystify Prognosis of Cancer of Unknown Primary (CUP) patients": All supplementary materials

Table S1

| | Som. mut. | CNA | Mut. sig. | | Som. mut. | CNA | Mut. sig. | | Som. mut. | CNA | Mut. sig. |
|---|---|---|---|---|---|---|---|---|---|---|---|
| ABL1 | ○ | ○ | | MYD88 | ○ | ○ | | GALNT12 | | ○ | |
| ACVR1 | ○ | ○ | | NBN | ○ | ○ | | GBA | | ○ | |
| AKT1 | ○ | ○ | | NEGR1 | ○ | ○ | | GEN1 | | ○ | |
| AKT2 | ○ | ○ | | NF1 | ○ | ○ | | GLI2 | | ○ | |
| AKT3 | ○ | ○ | | NF2 | ○ | ○ | | GLI3 | | ○ | |
| ALK | ○ | ○ | | NFE2L2 | ○ | ○ | | GPC3 | | ○ | |
| ALOX12B | ○ | ○ | | NFKBIA | ○ | ○ | | GSTM5 | | ○ | |
| APC | ○ | ○ | | NKX2-1 | ○ | ○ | | H19 | | ○ | |
| AR | ○ | ○ | | NKX3-1 | ○ | ○ | | HABP2 | | ○ | |
| ARAF | ○ | ○ | | NOTCH1 | ○ | ○ | | HELQ | | ○ | |
| ARID1A | ○ | ○ | | NOTCH2 | ○ | ○ | | HFE | | ○ | |
| ARID1B | ○ | ○ | | NOTCH3 | ○ | ○ | | HMBS | | ○ | |
| ARID2 | ○ | ○ | | NPM1 | ○ | ○ | | ID4 | | ○ | |
| ASXL1 | ○ | ○ | | NRAS | ○ | ○ | | IKZF3 | | ○ | |
| ATM | ○ | ○ | | NSD1 | ○ | ○ | | INSIG1 | | ○ | |
| ATR | ○ | ○ | | NTHL1 | ○ | ○ | | ITK | | ○ | |
| ATRX | ○ | ○ | | NTRK1 | ○ | ○ | | JAZF1 | | ○ | |
| AURKA | ○ | ○ | | NTRK2 | ○ | ○ | | KAT6B | | ○ | |
| AURKB | ○ | ○ | | NTRK3 | ○ | ○ | | KCNIP1 | | ○ | |
| AXIN2 | ○ | ○ | | PALB2 | ○ | ○ | | KCNQ1 | | ○ | |
| AXL | ○ | ○ | | PARK2 | ○ | ○ | | KDM6B | | ○ | |
| B2M | ○ | ○ | | PAX5 | ○ | ○ | | KIF1B | | ○ | |
| BABAM1 | ○ | ○ | | PBRM1 | ○ | ○ | | KLF2 | | ○ | |
| BAP1 | ○ | ○ | | PDCD1LG2 | ○ | ○ | | KLLN | | ○ | |
| BARD1 | ○ | ○ | | PDGFRA | ○ | ○ | | LIG4 | | ○ | |
| BCL2 | ○ | ○ | | PDGFRB | ○ | ○ | | LINC00894 | | ○ | |
| BCL2L1 | ○ | ○ | | PHOX2B | ○ | ○ | | LMO2 | | ○ | |
| BCL6 | ○ | ○ | | PIK3C2B | ○ | ○ | | LMO3 | | ○ | |
| BCOR | ○ | ○ | | PIK3CA | ○ | ○ | | MAF | | ○ | |
| BCORL1 | ○ | ○ | | PIK3R1 | ○ | ○ | | MAFB | | ○ | |
| BLM | ○ | ○ | | PIM1 | ○ | ○ | | MBD4 | | ○ | |
| BMPR1A | ○ | ○ | | PMS1 | ○ | ○ | | MCM8 | | ○ | |
| BRAF | ○ | ○ | | PMS2 | ○ | ○ | | MECOM | | ○ | |
| BRCA1 | ○ | ○ | | PNRC1 | ○ | ○ | | MLH3 | | ○ | |
| BRCA2 | ○ | ○ | | POLD1 | ○ | ○ | | MTA1 | | ○ | |
| BRD4 | ○ | ○ | | POLE | ○ | ○ | | MTAP | | ○ | |
| BRIP1 | ○ | ○ | | PPARG | ○ | ○ | | MUS81 | | ○ | |
| CALR | ○ | ○ | | PPM1D | ○ | ○ | | MYB | | ○ | |
| CARD11 | ○ | ○ | | PPP2R1A | ○ | ○ | | MYBL1 | | ○ | |
| CASP8 | ○ | ○ | | PRDM1 | ○ | ○ | | NEIL1 | | ○ | |
| CBFB | ○ | ○ | | PRKAR1A | ○ | ○ | | NEIL2 | | ○ | |
| CBL | ○ | ○ | | PRKCI | ○ | ○ | | NEIL3 | | ○ | |
| CCND1 | ○ | ○ | | PRKDC | ○ | ○ | | NFKBIE | | ○ | |
| CCND2 | ○ | ○ | | PTCH1 | ○ | ○ | | NFKBIZ | | ○ | |

**Table S1 continued from previous page**

| | | | | | | | | |
|---|---|---|---|---|---|---|---|---|
| CCND3 | ○ | ○ | PTEN | ○ | ○ | NPRL2 | | ○ |
| CCNE1 | ○ | ○ | PTPN11 | ○ | ○ | NPRL3 | | ○ |
| CD274 | ○ | ○ | PTPRD | ○ | ○ | NR0B1 | | ○ |
| CD79B | ○ | ○ | QKI | ○ | ○ | NRG1 | | ○ |
| CDC73 | ○ | ○ | RAC1 | ○ | ○ | NT5C2 | | ○ |
| CDH1 | ○ | ○ | RAD21 | ○ | ○ | OGG1 | | ○ |
| CDK12 | ○ | ○ | RAD50 | ○ | ○ | PAXIP1 | | ○ |
| CDK4 | ○ | ○ | RAD51 | ○ | ○ | PHF6 | | ○ |
| CDK6 | ○ | ○ | RAD51C | ○ | ○ | PML | | ○ |
| CDK8 | ○ | ○ | RAD51D | ○ | ○ | PNKP | | ○ |
| CDKN1A | ○ | ○ | RAD52 | ○ | ○ | POLB | | ○ |
| CDKN1B | ○ | ○ | RAF1 | ○ | ○ | POLH | | ○ |
| CDKN2A | ○ | ○ | RARA | ○ | ○ | POLQ | | ○ |
| CDKN2B | ○ | ○ | RASA1 | ○ | ○ | POT1 | | ○ |
| CDKN2C | ○ | ○ | RB1 | ○ | ○ | PRAME | | ○ |
| CEBPA | ○ | ○ | RBM10 | ○ | ○ | PRF1 | | ○ |
| CHEK1 | ○ | ○ | RECQL4 | ○ | ○ | PRKCZ | | ○ |
| CHEK2 | ○ | ○ | REL | ○ | ○ | PRPF40B | | ○ |
| CIC | ○ | ○ | RET | ○ | ○ | PRPF8 | | ○ |
| CREBBP | ○ | ○ | RFWD2 | ○ | ○ | PRSS1 | | ○ |
| CRKL | ○ | ○ | RHEB | ○ | ○ | PSMD13 | | ○ |
| CRLF2 | ○ | ○ | RHOA | ○ | ○ | PTK2 | | ○ |
| CSF1R | ○ | ○ | RICTOR | ○ | ○ | PTK2B | | ○ |
| CSF3R | ○ | ○ | RIT1 | ○ | ○ | PTPN14 | | ○ |
| CTCF | ○ | ○ | RNF43 | ○ | ○ | PVRL4 | | ○ |
| CTLA4 | ○ | ○ | ROS1 | ○ | ○ | RAD54B | | ○ |
| CTNNA1 | ○ | ○ | RPTOR | ○ | ○ | RBBP8 | | ○ |
| CTNNB1 | ○ | ○ | RUNX1 | ○ | ○ | RBL2 | | ○ |
| CXCR4 | ○ | ○ | RUNX1T1 | ○ | ○ | RELA | | ○ |
| CYLD | ○ | ○ | SDHA | ○ | ○ | RHBDF2 | | ○ |
| DAXX | ○ | ○ | SDHAF2 | ○ | ○ | RHOH | | ○ |
| DDR2 | ○ | ○ | SDHB | ○ | ○ | RHOT1 | | ○ |
| DICER1 | ○ | ○ | SDHC | ○ | ○ | RHPN2 | | ○ |
| DIS3 | ○ | ○ | SDHD | ○ | ○ | RIF1 | | ○ |
| DNMT3A | ○ | ○ | SETD2 | ○ | ○ | RINT1 | | ○ |
| EED | ○ | ○ | SF3B1 | ○ | ○ | RNF8 | | ○ |
| EGFR | ○ | ○ | SH2B3 | ○ | ○ | RPA1 | | ○ |
| EP300 | ○ | ○ | SH2D1A | ○ | ○ | RPL26 | | ○ |
| EPCAM | ○ | ○ | SLX4 | ○ | ○ | RSPO2 | | ○ |
| EPHA3 | ○ | ○ | SMAD2 | ○ | ○ | RSPO3 | | ○ |
| EPHA5 | ○ | ○ | SMAD4 | ○ | ○ | SBDS | | ○ |
| EPHA7 | ○ | ○ | SMARCA4 | ○ | ○ | SERPINA1 | | ○ |
| ERBB2 | ○ | ○ | SMARCB1 | ○ | ○ | SETBP1 | | ○ |
| ERBB3 | ○ | ○ | SMO | ○ | ○ | SF1 | | ○ |
| ERBB4 | ○ | ○ | SOCS1 | ○ | ○ | SLC25A13 | | ○ |
| ERCC2 | ○ | ○ | SOS1 | ○ | ○ | SLC34A2 | | ○ |
| ERCC3 | ○ | ○ | SOX2 | ○ | ○ | SLITRK6 | | ○ |
| ERCC4 | ○ | ○ | SOX9 | ○ | ○ | SLX1A | | ○ |
| ERCC5 | ○ | ○ | SPOP | ○ | ○ | SLX1B | | ○ |
| ERG | ○ | ○ | SRC | ○ | ○ | SMARCE1 | | ○ |

| | | | | | | | | |
|---|---|---|---|---|---|---|---|---|
| ESR1 | ○ | ○ | SRSF2 | ○ | ○ | SMC1A | ○ | |
| ETV1 | ○ | ○ | STAG2 | ○ | ○ | SMC3 | ○ | |
| ETV6 | ○ | ○ | STAT3 | ○ | ○ | SQSTM1 | ○ | |
| EZH2 | ○ | ○ | STK11 | ○ | ○ | SS18 | ○ | |
| FAM175A | ○ | ○ | SUFU | ○ | ○ | STAG1 | ○ | |
| FAM46C | ○ | ○ | SUZ12 | ○ | ○ | STAT6 | ○ | |
| FANCA | ○ | ○ | SYK | ○ | ○ | TAL1 | ○ | |
| FANCC | ○ | ○ | TCEB1 | ○ | ○ | TAL2 | ○ | |
| FANCD2 | ○ | ○ | TCF3 | ○ | ○ | TAZ | ○ | |
| FANCE | ○ | ○ | TCF7L2 | ○ | ○ | TCF7L1 | ○ | |
| FANCF | ○ | ○ | TERT | ○ | ○ | TDG | ○ | |
| FANCG | ○ | ○ | TET1 | ○ | ○ | TERC | ○ | |
| FANCL | ○ | ○ | TET2 | ○ | ○ | TFE3 | ○ | |
| FAS | ○ | ○ | TMEM127 | ○ | ○ | TLR4 | ○ | |
| FAT1 | ○ | ○ | TMPRSS2 | ○ | ○ | TLX3 | ○ | |
| FBXW7 | ○ | ○ | TNFAIP3 | ○ | ○ | TOPBP1 | ○ | |
| FGFR1 | ○ | ○ | TP53 | ○ | ○ | TRAF3 | ○ | |
| FGFR2 | ○ | ○ | TP53BP1 | ○ | ○ | TRIM37 | ○ | |
| FGFR3 | ○ | ○ | TRAF7 | ○ | ○ | UBE2T | ○ | |
| FGFR4 | ○ | ○ | TSC1 | ○ | ○ | UIMC1 | ○ | |
| FH | ○ | ○ | TSC2 | ○ | ○ | UROD | ○ | |
| FLCN | ○ | ○ | TSHR | ○ | ○ | USP28 | ○ | |
| FLT1 | ○ | ○ | U2AF1 | ○ | ○ | USP8 | ○ | |
| FLT3 | ○ | ○ | VEGFA | ○ | ○ | WAS | ○ | |
| FLT4 | ○ | ○ | VHL | ○ | ○ | WRN | ○ | |
| FOXA1 | ○ | ○ | WHSC1 | ○ | ○ | XPA | ○ | |
| FOXL2 | ○ | ○ | WHSC1L1 | ○ | ○ | XPC | ○ | |
| GATA2 | ○ | ○ | WT1 | ○ | ○ | XRCC1 | ○ | |
| GATA3 | ○ | ○ | XPO1 | ○ | ○ | XRCC3 | ○ | |
| GATA4 | ○ | ○ | XRCC2 | ○ | ○ | XRCC4 | ○ | |
| GATA6 | ○ | ○ | YAP1 | ○ | ○ | XRCC5 | ○ | |
| GLI1 | ○ | ○ | ZNF217 | ○ | ○ | XRCC6 | ○ | |
| GNA11 | ○ | ○ | ZRSR2 | ○ | ○ | ZNF708 | ○ | |
| GNAQ | ○ | ○ | BIRC3 | ○ | | ZNRF3 | ○ | |
| GNAS | ○ | ○ | FOXO1 | ○ | | SBS1 | | ○ |
| GREM1 | ○ | ○ | INSRR | ○ | | SBS2 | | ○ |
| H3F3A | ○ | ○ | MFSD11 | ○ | | SBS3 | | ○ |
| H3F3B | ○ | ○ | WWTR1 | ○ | | SBS4 | | ○ |
| HIST1H3B | ○ | ○ | ABCB11 | | ○ | SBS5 | | ○ |
| HIST1H3C | ○ | ○ | ARHGAP35 | | ○ | SBS6 | | ○ |
| HNF1A | ○ | ○ | ARHGEF12 | | ○ | SBS7a | | ○ |
| HOXB13 | ○ | ○ | BCL11B | | ○ | SBS7b | | ○ |
| HRAS | ○ | ○ | BCL2L12 | | ○ | SBS7c | | ○ |
| ID3 | ○ | ○ | BRCC3 | | ○ | SBS7d | | ○ |
| IDH1 | ○ | ○ | BRD3 | | ○ | SBS8 | | ○ |
| IDH2 | ○ | ○ | BRE | | ○ | SBS9 | | ○ |
| IGF1R | ○ | ○ | BUB1B | | ○ | SBS10a | | ○ |
| IGF2 | ○ | ○ | CADM2 | | ○ | SBS10b | | ○ |
| IKZF1 | ○ | ○ | CBFA2T3 | | ○ | SBS11 | | ○ |
| IL7R | ○ | ○ | CBLB | | ○ | SBS12 | | ○ |

**Table S1 continued from previous page**

| | | | | | | |
|---|---|---|---|---|---|---|
| JAK1 | ○ | ○ | CD58 | ○ | SBS13 | ○ |
| JAK2 | ○ | ○ | CDH4 | ○ | SBS14 | ○ |
| JAK3 | ○ | ○ | CDK1 | ○ | SBS15 | ○ |
| KAT6A | ○ | ○ | CDK2 | ○ | SBS16 | ○ |
| KDM5A | ○ | ○ | CDK5 | ○ | SBS17a | ○ |
| KDM5C | ○ | ○ | CDK9 | ○ | SBS17b | ○ |
| KDM6A | ○ | ○ | CDKN1C | ○ | SBS18 | ○ |
| KDR | ○ | ○ | CIITA | ○ | SBS19 | ○ |
| KEAP1 | ○ | ○ | COL7A1 | ○ | SBS20 | ○ |
| KIT | ○ | ○ | CRTC1 | ○ | SBS21 | ○ |
| KLF4 | ○ | ○ | CRTC2 | ○ | SBS22 | ○ |
| KMT2A | ○ | ○ | CUX1 | ○ | SBS23 | ○ |
| KMT2D | ○ | ○ | DCLRE1C | ○ | SBS24 | ○ |
| KRAS | ○ | ○ | DDB1 | ○ | SBS25 | ○ |
| LMO1 | ○ | ○ | DDB2 | ○ | SBS26 | ○ |
| MAP2K1 | ○ | ○ | DEPDC5 | ○ | SBS28 | ○ |
| MAP2K2 | ○ | ○ | DIS3L2 | ○ | SBS29 | ○ |
| MAP2K4 | ○ | ○ | DKC1 | ○ | SBS30 | ○ |
| MAP3K1 | ○ | ○ | DMC1 | ○ | SBS31 | ○ |
| MAPK1 | ○ | ○ | DMD | ○ | SBS32 | ○ |
| MAX | ○ | ○ | DOCK8 | ○ | SBS33 | ○ |
| MCL1 | ○ | ○ | EGLN1 | ○ | SBS34 | ○ |
| MDM2 | ○ | ○ | ELANE | ○ | SBS35 | ○ |
| MDM4 | ○ | ○ | EME1 | ○ | SBS36 | ○ |
| MED12 | ○ | ○ | ENG | ○ | SBS37 | ○ |
| MEF2B | ○ | ○ | ERCC1 | ○ | SBS38 | ○ |
| MEN1 | ○ | ○ | ERCC6 | ○ | SBS39 | ○ |
| MET | ○ | ○ | ETV4 | ○ | SBS40 | ○ |
| MGA | ○ | ○ | ETV5 | ○ | SBS41 | ○ |
| MITF | ○ | ○ | EWSR1 | ○ | SBS42 | ○ |
| MLH1 | ○ | ○ | EXO1 | ○ | SBS44 | ○ |
| MPL | ○ | ○ | EXT1 | ○ | SBS84 | ○ |
| MRE11A | ○ | ○ | EXT2 | ○ | SBS85 | ○ |
| MSH2 | ○ | ○ | FAH | ○ | SBS86 | ○ |
| MSH6 | ○ | ○ | FAN1 | ○ | SBS87 | ○ |
| MTOR | ○ | ○ | FANCB | ○ | SBS88 | ○ |
| MUTYH | ○ | ○ | FANCI | ○ | SBS89 | ○ |
| MYC | ○ | ○ | FANCM | ○ | SBS90 | ○ |
| MYCL | ○ | ○ | FKBP9 | ○ | | |
| MYCN | ○ | ○ | FUS | ○ | | |

### Table S2

| | | Minimum $p_{\max}$ threshold | | | |
|---|---|---|---|---|---|
| | | 0.0 | 0.5 | 0.7 | 0.9 |
| Overall | DFCI | 3690 | 3438 | 3047 | 2502 |
| | MSK | 3331 | 3012 | 2608 | 2112 |
| | VICC | 268 | 230 | 192 | 136 |
| Non-Small Cell Lung Cancer (NSCLC) | DFCI | 811 | 735 | 644 | 533 |
| | MSK | 717 | 618 | 520 | 430 |
| | VICC | 36 | 27 | 23 | 19 |
| Invasive Breast Carcinoma (BRCA) | DFCI | 600 | 572 | 514 | 433 |
| | MSK | 727 | 675 | 598 | 474 |
| | VICC | 68 | 62 | 48 | 35 |
| Colorectal Adenocarcinoma (COADREAD) | DFCI | 521 | 502 | 479 | 436 |
| | MSK | 375 | 358 | 330 | 303 |
| | VICC | 55 | 52 | 48 | 37 |
| Diffuse Glioma (DIFG) | DFCI | 400 | 390 | 383 | 361 |
| | MSK | 214 | 204 | 187 | 168 |
| | VICC | 11 | 10 | 8 | 4 |
| Prostate Adenocarcinoma (PRAD) | DFCI | 126 | 118 | 98 | 67 |
| | MSK | 300 | 280 | 233 | 163 |
| | VICC | 16 | 10 | 6 | 3 |
| Pancreatic Adenocarcinoma (PAAD) | DFCI | 136 | 125 | 104 | 71 |
| | MSK | 233 | 216 | 187 | 154 |
| | VICC | 10 | 8 | 6 | 1 |
| Ovarian Epithelial Tumor (OVT) | DFCI | 257 | 229 | 184 | 112 |
| | MSK | 100 | 60 | 38 | 10 |
| | VICC | 12 | 9 | 5 | 2 |
| Esophagogastric Adenocarcinoma (EGC) | DFCI | 171 | 153 | 114 | 66 |
| | MSK | 82 | 70 | 44 | 24 |
| | VICC | 11 | 8 | 7 | 2 |
| Endometrial Carcinoma (UCEC) | DFCI | 123 | 116 | 95 | 73 |
| | MSK | 105 | 100 | 91 | 70 |
| | VICC | 7 | 6 | 6 | 2 |
| Melanoma (MEL) | DFCI | 134 | 127 | 115 | 103 |
| | MSK | 108 | 103 | 98 | 92 |
| | VICC | 24 | 24 | 23 | 23 |
| Bladder Urothelial Carcinoma (BLCA) | DFCI | 86 | 81 | 67 | 52 |
| | MSK | 93 | 84 | 78 | 65 |
| | VICC | 4 | 4 | 4 | 3 |

| | | Minimum $p_{\max}$ threshold | | | |
|---|---|---|---|---|---|
| | | 0.0 | 0.5 | 0.7 | 0.9 |
| Renal Cell Carcinoma (RCC) | DFCI | 79 | 71 | 61 | 50 |
| | MSK | 85 | 75 | 68 | 56 |
| | VICC | 6 | 5 | 4 | 3 |
| Head and Neck Squamous Cell Carcinoma (HNSCC) | DFCI | 55 | 50 | 39 | 28 |
| | MSK | 27 | 18 | 12 | 5 |
| | VICC | . | . | . | . |
| Cholangiocarcinoma (CHOL) | DFCI | 18 | 12 | 10 | 7 |
| | MSK | 40 | 31 | 24 | 16 |
| | VICC | 1 | . | . | . |
| Gastrointestinal Stromal Tumor (GIST) | DFCI | 47 | 46 | 43 | 40 |
| | MSK | 34 | 33 | 31 | 30 |
| | VICC | . | . | . | . |
| Well-Differentiated Thyroid Cancer (WDTC) | DFCI | 17 | 15 | 14 | 9 |
| | MSK | 31 | 31 | 29 | 25 |
| | VICC | 1 | 1 | 1 | . |
| Pleural Mesothelioma (PLMESO) | DFCI | 24 | 21 | 14 | 10 |
| | MSK | 18 | 17 | 10 | 6 |
| | VICC | 5 | 3 | 2 | 1 |
| Meningothelial Tumor (MNGT) | DFCI | 27 | 25 | 23 | 20 |
| | MSK | 3 | 3 | 1 | . |
| | VICC | 1 | 1 | 1 | 1 |
| Gastrointestinal Neuroendocrine Tumors (GINET) | DFCI | 20 | 17 | 16 | 11 |
| | MSK | 3 | 3 | 2 | . |
| | VICC | . | . | . | . |
| Pancreatic Neuroendocrine Tumor (PANET) | DFCI | 15 | 14 | 13 | 8 |
| | MSK | 24 | 22 | 19 | 15 |
| | VICC | . | . | . | . |
| Acute Myeloid Leukemia (AML) | DFCI | 15 | 11 | 10 | 6 |
| | MSK | . | . | . | . |
| | VICC | . | . | . | . |
| Non-Hodgkin Lymphoma (NHL) | DFCI | 8 | 8 | 7 | 6 |
| | MSK | 12 | 11 | 8 | 6 |
| | VICC | . | . | . | . |

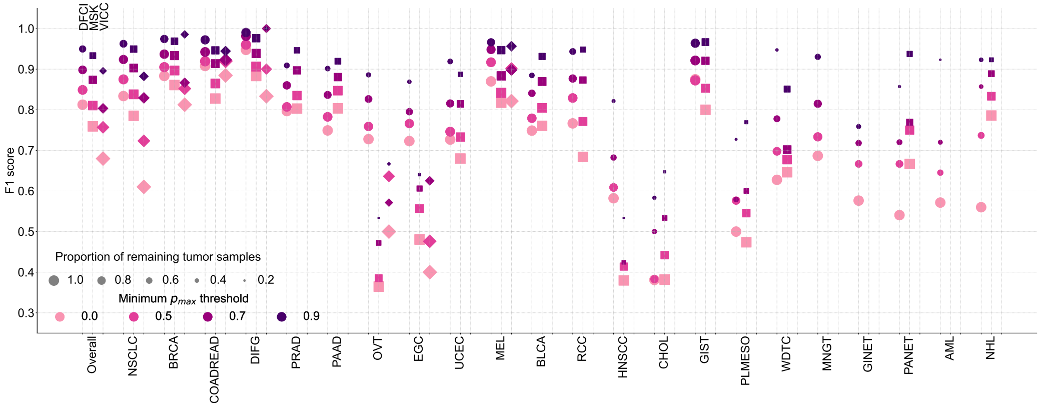


(a)

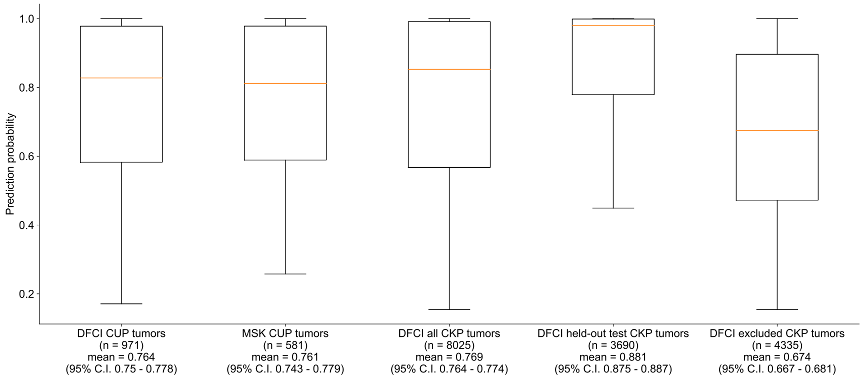


(b)

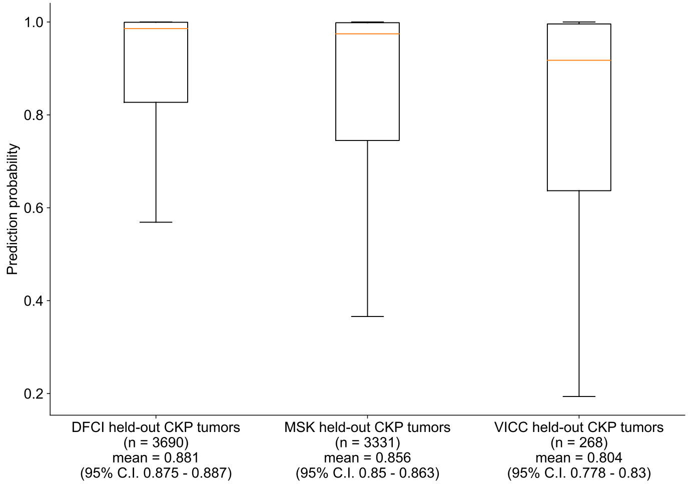


(c)

Figure S1

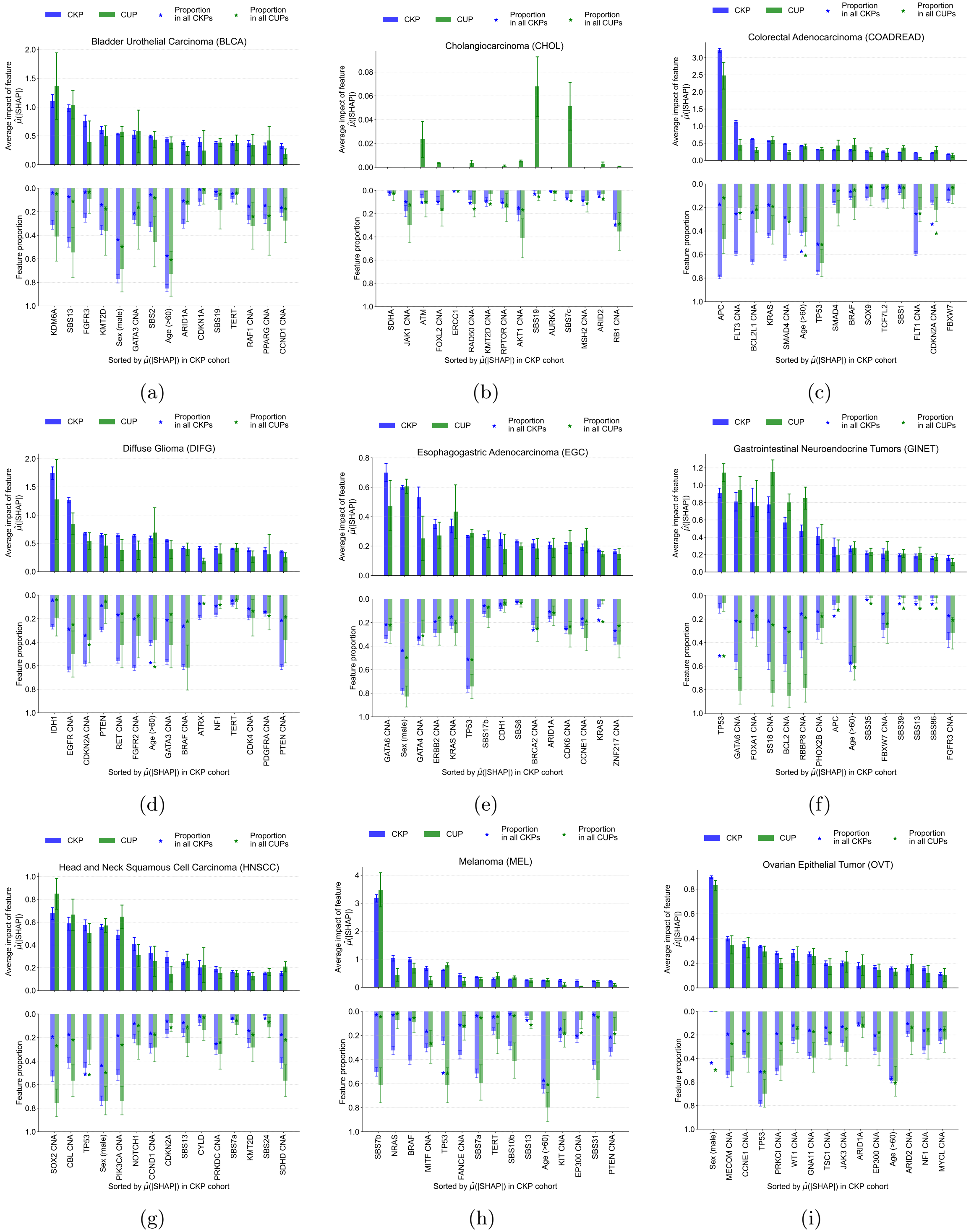
CKP
CUP
Proportion in all CKPs
Proportion in all CUPs
Average impact of feature $\hat{\mu}$(|SHAP|)
Feature proportion
Sorted by $\hat{\mu}$(|SHAP|) in CKP cohort
Bladder Urothelial Carcinoma (BLCA)
KDM6A
SBS13
FGFR3
KMT2D
Sex (male)
GATA3 CNA
SBS2
Age (>60)
ARID1A
CDKN1A
SBS19
TERT
RAF1 CNA
PPARG CNA
CCND1 CNA
(a)
Cholangiocarcinoma (CHOL)
SDHA
JAK1 CNA
ATM
FOXL2 CNA
ERCC1
RAD50 CNA
KMT2D CNA
RPTOR CNA
AKT1 CNA
SBS19
AURKA
SBS7c
MSH2 CNA
ARID2
RB1 CNA
(b)
Colorectal Adenocarcinoma (COADREAD)
APC
FLT3 CNA
BCL2L1 CNA
KRAS
SMAD4 CNA
Age (>60)
TP53
SMAD4
BRAF
SOX9
TCF7L2
SBS1
FLT1 CNA
CDKN2A CNA
FBXW7
(c)
Diffuse Glioma (DIFG)
IDH1
EGFR CNA
CDKN2A CNA
PTEN
RET CNA
FGFR2 CNA
Age (>60)
GATA3 CNA
BRAF CNA
ATRX
NF1
TERT
CDK4 CNA
PDGFRA CNA
PTEN CNA
(d)
Esophagogastric Adenocarcinoma (EGC)
GATA6 CNA
Sex (male)
GATA4 CNA
ERBB2 CNA
KRAS CNA
TP53
SBS17b
CDH1
SBS6
BRCA2 CNA
ARID1A
CDK6 CNA
CCNE1 CNA
KRAS
ZNF217 CNA
(e)
Gastrointestinal Neuroendocrine Tumors (GINET)
TP53
GATA6 CNA
FOXA1 CNA
SS18 CNA
BCL2 CNA
RBBP8 CNA
PHOX2B CNA
APC
Age (>60)
SBS35
FBXW7 CNA
SBS39
SBS13
SBS86
FGFR3 CNA
(f)
Head and Neck Squamous Cell Carcinoma (HNSCC)
SOX2 CNA
CBL CNA
TP53
Sex (male)
PIK3CA CNA
NOTCH1
CCND1 CNA
CDKN2A
SBS13
CYLD
PRKDC CNA
SBS7a
KMT2D
SBS24
SDHD CNA
(g)
Melanoma (MEL)
SBS7b
NRAS
BRAF
MITF CNA
TP53
FANCE CNA
SBS7a
TERT
SBS10b
SBS13
Age (>60)
KIT CNA
EP300 CNA
SBS31
PTEN CNA
(h)
Ovarian Epithelial Tumor (OVT)
Sex (male)
MECOM CNA
CCNE1 CNA
TP53
PRKCI CNA
WT1 CNA
GNA11 CNA
TSC1 CNA
JAK3 CNA
ARID1A
EP300 CNA
Age (>60)
ARID2 CNA
NF1 CNA
MYCL CNA
(i)

Figure S2

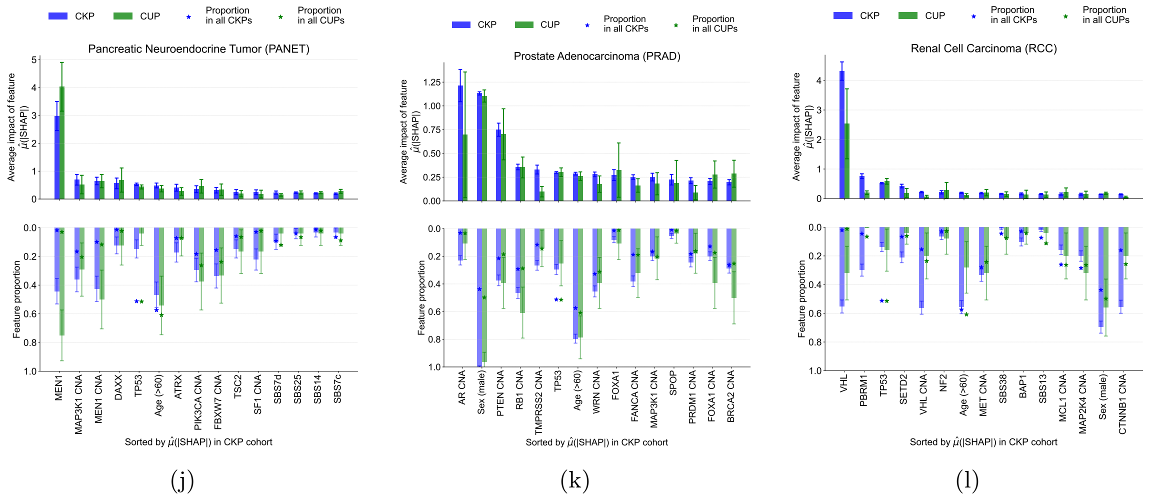

CKP
CUP
Proportion in all CKPs
Proportion in all CUPs
Pancreatic Neuroendocrine Tumor (PANET)
Average impact of feature $\hat{\mu}$(|SHAP|)
Feature proportion
MEN1
MAP3K1 CNA
MEN1 CNA
DAXX
TP53
Age (>60)
ATRX
PIK3CA CNA
FBXW7 CNA
TSC2
SF1 CNA
SBS7d
SBS25
SBS14
SBS7c
Sorted by $\hat{\mu}$(|SHAP|) in CKP cohort
(j)
Prostate Adenocarcinoma (PRAD)
AR CNA
Sex (male)
PTEN CNA
RB1 CNA
TMPRSS2 CNA
TP53
Age (>60)
WRN CNA
FOXA1
FANCA CNA
MAP3K1 CNA
SPOP
PRDM1 CNA
FOXA1 CNA
BRCA2 CNA
(k)
Renal Cell Carcinoma (RCC)
VHL
PBRM1
TP53
SETD2
VHL CNA
NF2
Age (>60)
MET CNA
SBS38
BAP1
SBS13
MCL1 CNA
MAP2K4 CNA
Sex (male)
CTNNB1 CNA
(l)


Figure S2

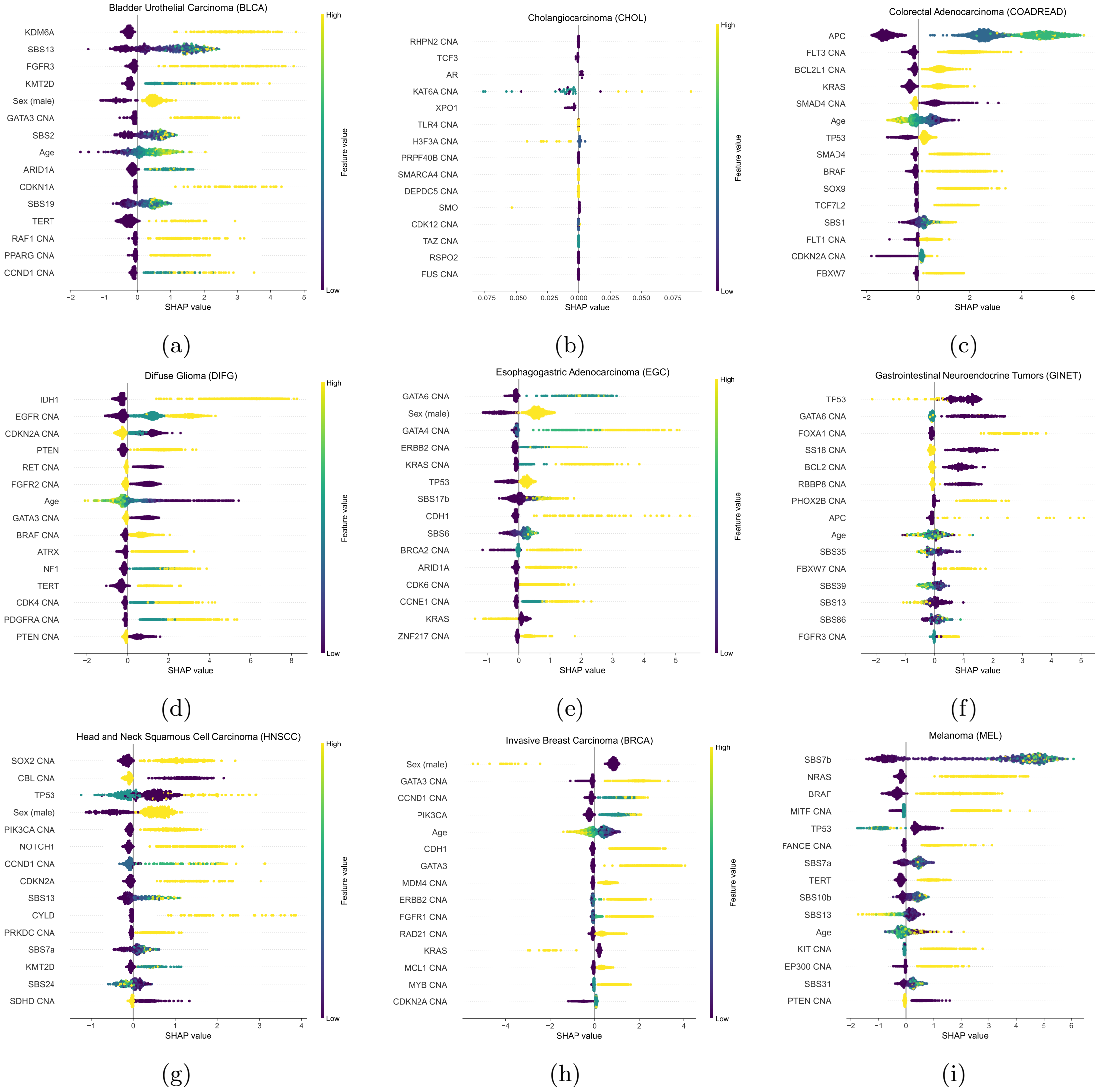

Bladder Urothelial Carcinoma (BLCA)
KDM6A
SBS13
FGFR3
KMT2D
Sex (male)
GATA3 CNA
SBS2
Age
ARID1A
CDKN1A
SBS19
TERT
RAF1 CNA
PPARG CNA
CCND1 CNA
SHAP value
Feature value
High
Low
(a)
Cholangiocarcinoma (CHOL)
RHPN2 CNA
TCF3
AR
KAT6A CNA
XPO1
TLR4 CNA
H3F3A CNA
PRPF40B CNA
SMARCA4 CNA
DEPDC5 CNA
SMO
CDK12 CNA
TAZ CNA
RSPO2
FUS CNA
SHAP value
Feature value
High
Low
(b)
Colorectal Adenocarcinoma (COADREAD)
APC
FLT3 CNA
BCL2L1 CNA
KRAS
SMAD4 CNA
Age
TP53
SMAD4
BRAF
SOX9
TCF7L2
SBS1
FLT1 CNA
CDKN2A CNA
FBXW7
SHAP value
Feature value
High
Low
(c)
Diffuse Glioma (DIFG)
IDH1
EGFR CNA
CDKN2A CNA
PTEN
RET CNA
FGFR2 CNA
Age
GATA3 CNA
BRAF CNA
ATRX
NF1
TERT
CDK4 CNA
PDGFRA CNA
PTEN CNA
SHAP value
Feature value
High
Low
(d)
Esophagogastric Adenocarcinoma (EGC)
GATA6 CNA
Sex (male)
GATA4 CNA
ERBB2 CNA
KRAS CNA
TP53
SBS17b
CDH1
SBS6
BRCA2 CNA
ARID1A
CDK6 CNA
CCNE1 CNA
KRAS
ZNF217 CNA
SHAP value
Feature value
High
Low
(e)
Gastrointestinal Neuroendocrine Tumors (GINET)
TP53
GATA6 CNA
FOXA1 CNA
SS18 CNA
BCL2 CNA
RBBP8 CNA
PHOX2B CNA
APC
Age
SBS35
FBXW7 CNA
SBS39
SBS13
SBS86
FGFR3 CNA
SHAP value
Feature value
High
Low
(f)
Head and Neck Squamous Cell Carcinoma (HNSCC)
SOX2 CNA
CBL CNA
TP53
Sex (male)
PIK3CA CNA
NOTCH1
CCND1 CNA
CDKN2A
SBS13
CYLD
PRKDC CNA
SBS7a
KMT2D
SBS24
SDHD CNA
SHAP value
Feature value
High
Low
(g)
Invasive Breast Carcinoma (BRCA)
Sex (male)
GATA3 CNA
CCND1 CNA
PIK3CA
Age
CDH1
GATA3
MDM4 CNA
ERBB2 CNA
FGFR1 CNA
RAD21 CNA
KRAS
MCL1 CNA
MYB CNA
CDKN2A CNA
SHAP value
Feature value
High
Low
(h)
Melanoma (MEL)
SBS7b
NRAS
BRAF
MITF CNA
TP53
FANCE CNA
SBS7a
TERT
SBS10b
SBS13
Age
KIT CNA
EP300 CNA
SBS31
PTEN CNA
SHAP value
Feature value
High
Low
(i)


Figure S3

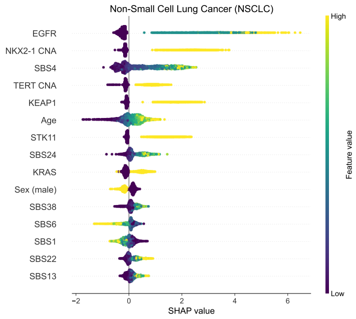


(j)

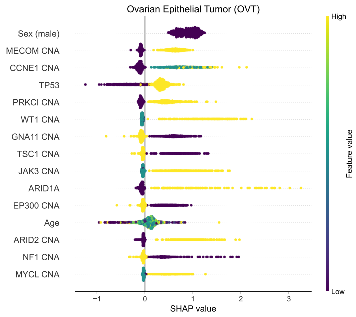


(k)

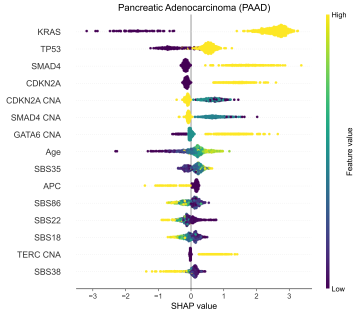


(l)

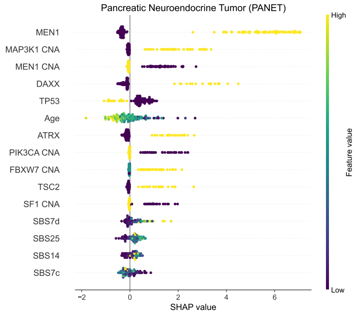


(m)

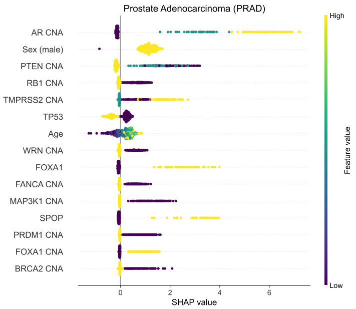


(n)

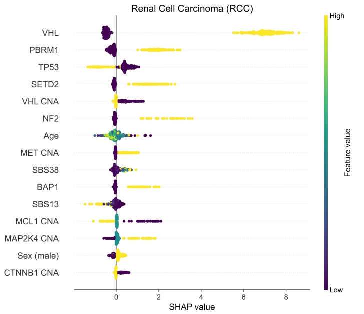


(o)

Figure S3

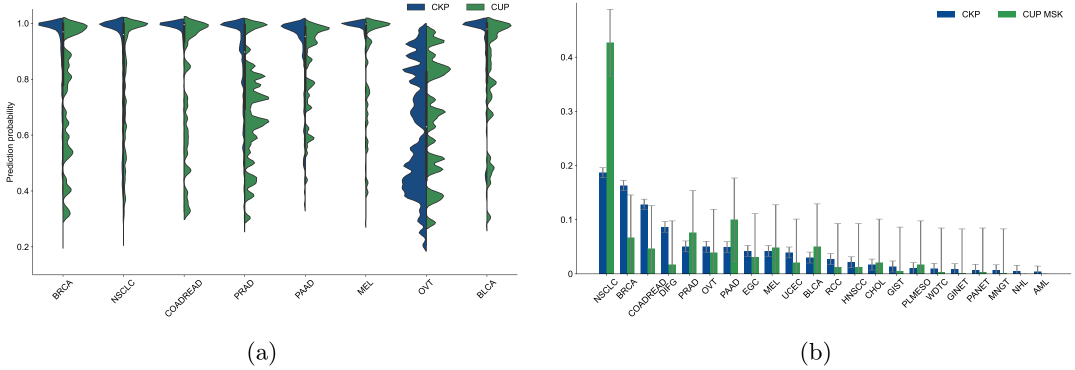
CKP
CUP
Prediction probability
1.0
0.8
0.6
0.4
0.2
BRCA
NSCLC
COADREAD
PRAD
PAAD
MEL
OVT
BLCA
(a)
CKP
CUP MSK
0.4
0.3
0.2
0.1
0
NSCLC
BRCA
COADREAD
DIFG
PRAD
OVT
PAAD
EGC
MEL
UCEC
BLCA
RCC
HNSCC
CHOL
GIST
PLMESO
WDTC
GINET
PANET
MNGT
NHL
AML
(b)

Figure S4

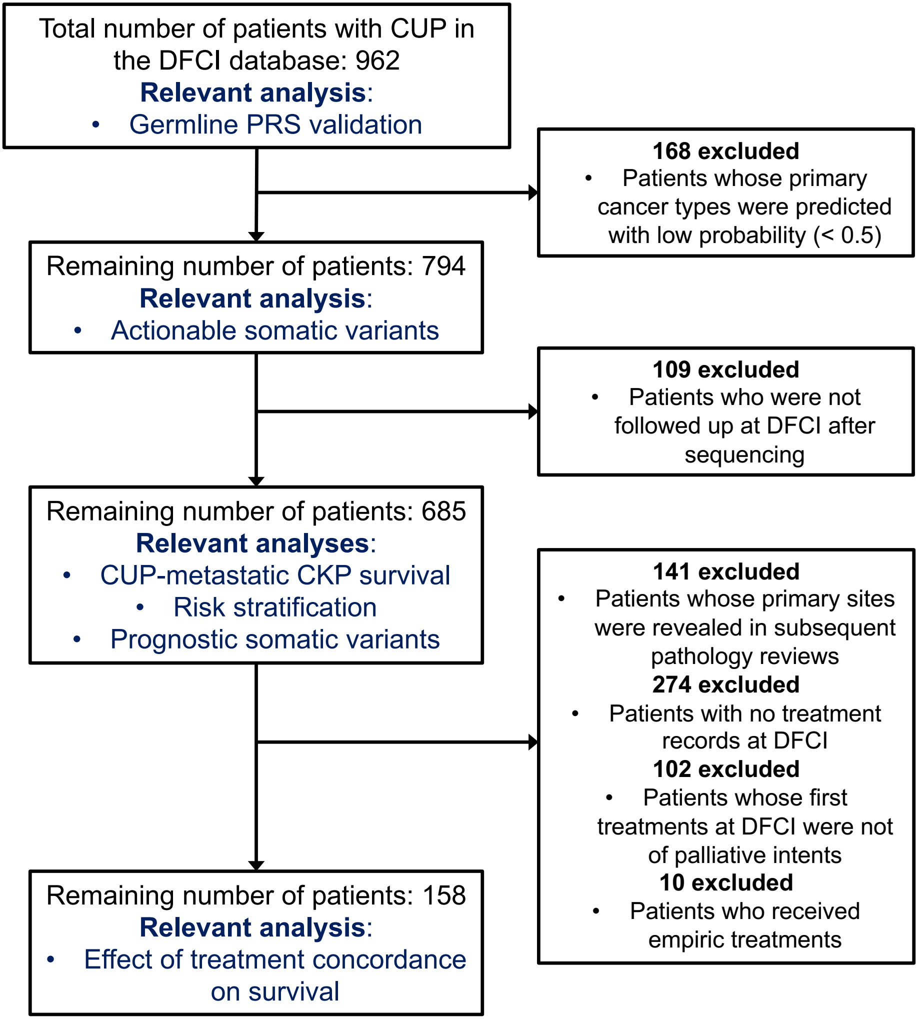
Total number of patients with CUP in the DFCI database: 962
Relevant analysis:
• Germline PRS validation
168 excluded
• Patients whose primary cancer types were predicted with low probability (< 0.5)
Remaining number of patients: 794
Relevant analysis:
• Actionable somatic variants
109 excluded
• Patients who were not followed up at DFCI after sequencing
Remaining number of patients: 685
Relevant analyses:
• CUP-metastatic CKP survival
• Risk stratification
• Prognostic somatic variants
141 excluded
• Patients whose primary sites were revealed in subsequent pathology reviews
274 excluded
• Patients with no treatment records at DFCI
102 excluded
• Patients whose first treatments at DFCI were not of palliative intents
10 excluded
• Patients who received empiric treatments
Remaining number of patients: 158
Relevant analysis:
• Effect of treatment concordance on survival

Figure S5

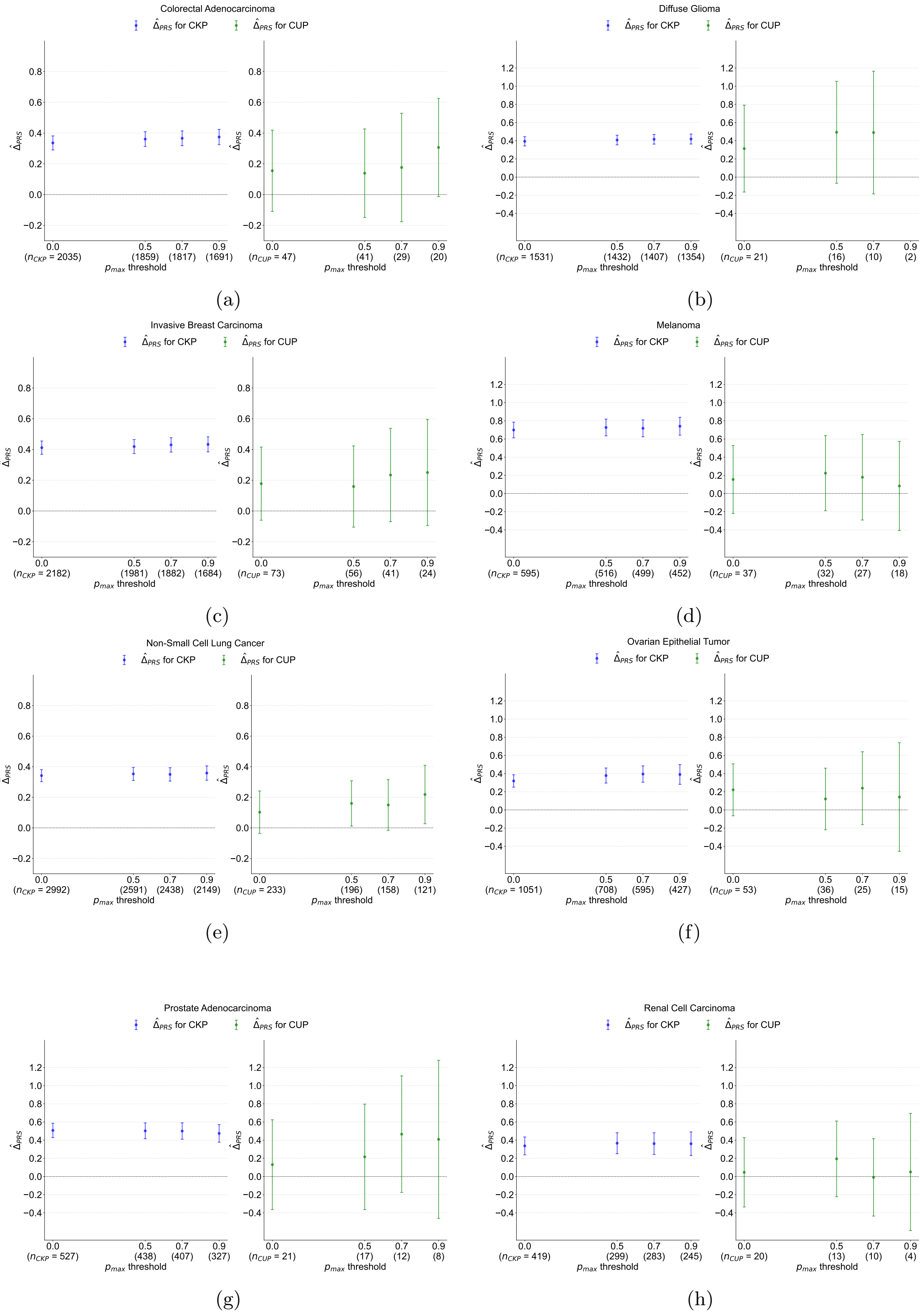
Colorectal Adenocarcinoma
$\hat{\Delta}_{PRS}$ for CKP
$\hat{\Delta}_{PRS}$ for CUP
($n_{CKP}$ = 2035) (1859) (1817) (1691)
($n_{CUP}$ = 47) (41) (29) (20)
(a)
Diffuse Glioma
($n_{CKP}$ = 1531) (1432) (1407) (1354)
($n_{CUP}$ = 21) (16) (10) (2)
(b)
Invasive Breast Carcinoma
($n_{CKP}$ = 2182) (1981) (1882) (1684)
($n_{CUP}$ = 73) (56) (41) (24)
(c)
Melanoma
($n_{CKP}$ = 595) (516) (499) (452)
($n_{CUP}$ = 37) (32) (27) (18)
(d)
Non-Small Cell Lung Cancer
($n_{CKP}$ = 2992) (2591) (2438) (2149)
($n_{CUP}$ = 233) (196) (158) (121)
(e)
Ovarian Epithelial Tumor
($n_{CKP}$ = 1051) (708) (595) (427)
($n_{CUP}$ = 53) (36) (25) (15)
(f)
Prostate Adenocarcinoma
($n_{CKP}$ = 527) (438) (407) (327)
($n_{CUP}$ = 21) (17) (12) (8)
(g)
Renal Cell Carcinoma
($n_{CKP}$ = 419) (299) (283) (245)
($n_{CUP}$ = 20) (13) (10) (4)
(h)
$p_{max}$ threshold
$\hat{\Delta}_{PRS}$

Figure S6

Table S3

| Pt. idx | Primary cancer diagnosis | Biopsy site | Treatment -OncoNPC concodrance | Sex | OncoNPC prediction | First chemo plan at DFCI | 5-year death |
|---|---|---|---|---|---|---|---|
| 0 | Poorly Differentiated Carcinoma, NOS | SOFT_TISSUE | FALSE | MALE | PRAD | CAPECITABINE | 1 |
| 1 | Adenocarcinoma, NOS | PERITONEUM | FALSE | FEMALE | BRCA | CARBOPLATIN/PACLITAXEL | 1 |
| 2 | Adenocarcinoma, NOS | LIVER | TRUE | MALE | PAAD | PACLITAXEL/GEMCITABINE | 1 |
| 3 | Neuroendocrine Tumor, NOS | LIVER | FALSE | MALE | NSCLC | ETOPOSIDE/CARBOPLATIN | 0 |
| 4 | Cancer of Unknown Primary | BRAIN | FALSE | MALE | NSCLC | ETOPOSIDE/IFOSFAMIDE/ CISPLATIN | 0 |
| 5 | Neuroendocrine Tumor, NOS | LIVER | TRUE | MALE | GINET | OCTREOTIDE | 0 |
| 6 | Adenocarcinoma, NOS | LIVER | TRUE | FEMALE | CHOL | GEMCITABINE/CISPLATIN | 1 |
| 7 | Neuroendocrine Carcinoma, NOS | LIVER | FALSE | FEMALE | BRCA | ETOPOSIDE/CISPLATIN | 0 |
| 8 | Poorly Differentiated Carcinoma, NOS | BOWEL | SOFT FALSE | MALE | MEL | DOCETAXEL/CISPLATIN | 0 |
| 9 | Squamous Cell Carcinoma, NOS | PERITONEUM | FALSE | FEMALE | NSCLC | GEMCITABINE/CISPLATIN | 1 |
| 10 | Cancer of Unknown Primary | BONE | TRUE | MALE | NSCLC | PEMETREXED/CARBOPLATIN | 1 |
| 11 | Adenocarcinoma, NOS | BRAIN | FALSE | MALE | NSCLC | BEVACIZUMAB/OXALIPLATIN /FLUOROURACIL/LEUCOVORIN | 1 |
| 12 | Adenocarcinoma, NOS | PLEURA | SOFT FALSE | MALE | PAAD | IRINOTECAN/CISPLATIN | 1 |
| 13 | Cancer of Unknown Primary | LIVER | TRUE | MALE | PAAD | OXALIPLATIN/FLUOROURACIL /LEUCOVORIN | 1 |
| 14 | Cancer of Unknown Primary | SOFT_TISSUE | FALSE | MALE | BLCA | VISMODEGIB | 0 |
| 15 | Undifferentiated Malignant Neoplasm | SKIN | SOFT FALSE | MALE | MEL | PEMBROLIZUMAB | 1 |
| 16 | Adenocarcinoma, NOS | LIVER | FALSE | FEMALE | PAAD | GEMCITABINE/CISPLATIN | 1 |

**Table S3 continued from previous page**

| | | | | | | | |
|---|---|---|---|---|---|---|---|
| 17 | Cancer of Unknown Primary | LUNG | TRUE | FEMALE | NSCLC | PEMETREXED/CARBOPLATIN | 1 |
| 18 | Small Cell Carcinoma of Unknown Primary | LIVER | FALSE | MALE | COADREAD | ETOPOSIDE/CISPLATIN | 1 |
| 19 | Adenocarcinoma, NOS | LYMPH | FALSE | MALE | COADREAD | PEMETREXED/CARBOPLATIN | 1 |
| 20 | Cancer of Unknown Primary | BOWEL | TRUE | FEMALE | OVT | PACLITAXEL | 1 |
| 21 | Adenocarcinoma, NOS | LUNG | TRUE | FEMALE | OVT | CARBOPLATIN/PACLITAXEL | 0 |
| 22 | Undifferentiated Malignant Neoplasm | SOFT_TISSUE | TRUE | FEMALE | MEL | PEMBROLIZUMAB | 1 |
| 23 | Adenocarcinoma, NOS | LIVER | FALSE | FEMALE | PANET | GEMCITABINE/CISPLATIN | 1 |
| 24 | Adenocarcinoma, NOS | LIVER | FALSE | MALE | MEL | GEMCITABINE/CISPLATIN | 1 |
| 25 | Adenocarcinoma, NOS | SOFT_TISSUE | FALSE | MALE | NSCLC | GEMCITABINE/CISPLATIN | 0 |
| 26 | Cancer of Unknown Primary | PLEURA | FALSE | MALE | DIFG | CARBOPLATIN/PACLITAXEL | 1 |
| 27 | Adenocarcinoma, NOS | PLEURA | TRUE | FEMALE | OVT | CARBOPLATIN/PACLITAXEL | 0 |
| 28 | Adenocarcinoma, NOS | LIVER | FALSE | MALE | COADREAD | FOLFIRINOX | 1 |
| 29 | Poorly Differentiated Carcinoma, NOS | LIVER | TRUE | MALE | PAAD | PACLITAXEL/GEMCITABINE | 1 |
| 30 | Small Cell Carcinoma of Unknown Primary | LIVER | FALSE | FEMALE | BRCA | TOPOTECAN | 1 |
| 31 | Neuroendocrine Tumor, NOS | SKIN | FALSE | MALE | EGC | ETOPOSIDE/CISPLATIN | 1 |
| 32 | Neuroendocrine Tumor, NOS | LIVER | TRUE | FEMALE | BRCA | PACLITAXEL | 1 |
| 33 | Adenocarcinoma, NOS | BRAIN | FALSE | FEMALE | BRCA | FOLFIRI | 1 |
| 34 | Adenocarcinoma, NOS | SOFT_TISSUE | FALSE | FEMALE | NSCLC | OXALIPLATIN/FLUOROURACIL /LEUCOVORIN | 1 |
| 35 | Adenocarcinoma, NOS | LIVER | TRUE | FEMALE | PAAD | FOLFIRINOX | 1 |

**Table S3 continued from previous page**

| | | | | | | | |
|---|---|---|---|---|---|---|---|
| 36 | Cancer of Unknown Primary | LYMPH | TRUE | FEMALE | NSCLC | PEMETREXED/CARBOPLATIN | 1 |
| 37 | Adenocarcinoma, NOS | PLEURA | FALSE | FEMALE | BRCA | PEMBROLIZUMAB | 1 |
| 38 | Adenocarcinoma, NOS | SOFT_TISSUE | TRUE | FEMALE | OVT | CARBOPLATIN/GEMCITABINE | 1 |
| 39 | Poorly Differentiated Carcinoma, NOS | UNAVAILABL | FALSE | FEMALE | NSCLC | FLUOROURACIL/LEUCOVORIN | 1 |
| 40 | Adenocarcinoma, NOS | OVARY | TRUE | FEMALE | COADREAD | BEVACIZUMAB/OXALIPLATIN /FLUOROURACIL/LEUCOVORIN | 1 |
| 41 | Adenocarcinoma, NOS | LIVER | FALSE | MALE | NSCLC | GEMCITABINE/CISPLATIN | 1 |
| 42 | Cancer of Unknown Primary, NOS | SKIN | TRUE | FEMALE | BRCA | PERTUZUMAB/TRASTUZUMAB /PACLITAXEL | 0 |
| 43 | Squamous Cell Carcinoma, NOS | LIVER | FALSE | MALE | NSCLC | CAPECITABINE/OXALIPLATIN | 1 |
| 44 | Poorly Differentiated Carcinoma, NOS | LYMPH | TRUE | MALE | COADREAD | OXALIPLATIN/FLUOROURACIL /LEUCOVORIN | 0 |
| 45 | Undifferentiated Malignant Neoplasm | SOFT_TISSUE | FALSE | MALE | NSCLC | ETOPOSIDE/CISPLATIN | 1 |
| 46 | Adenocarcinoma, NOS | PERITONEUM | TRUE | FEMALE | PAAD | FOLFIRINOX | 1 |
| 47 | Adenocarcinoma, NOS | LUNG | FALSE | FEMALE | NSCLC | CARBOPLATIN/PACLITAXEL | 0 |
| 48 | Cancer of Unknown Primary, NOS | LUNG | TRUE | MALE | NSCLC | PEMETREXED/CARBOPLATIN | 1 |
| 49 | Neuroendocrine Tumor, NOS | LIVER | SOFT FALSE | MALE | PANET | EVEROLIMUS | 1 |
| 50 | Neuroendocrine Tumor, NOS | LIVER | FALSE | MALE | COADREAD | ETOPOSIDE/CISPLATIN | 1 |
| 51 | Neuroendocrine Tumor, NOS | BOWEL | TRUE | MALE | GINET | OCTREOTIDE | 0 |
| 52 | Poorly Differentiated Carcinoma, NOS | LYMPH | FALSE | FEMALE | BRCA | GEMCITABINE/CISPLATIN | 0 |
| 53 | Neuroendocrine Tumor, NOS | LYMPH | FALSE | FEMALE | NSCLC | IMATINIB | 0 |

**Table S3 continued from previous page**

| | | | | | | | |
|---|---|---|---|---|---|---|---|
| 54 | Poorly Differentiated Carcinoma, NOS | UTERUS | FALSE | FEMALE | GINET | CARBOPLATIN/PACLITAXEL | 1 |
| 55 | Poorly Differentiated Carcinoma, NOS | BLADDER | FALSE | MALE | PRAD | GEMCITABINE/CISPLATIN | 1 |
| 56 | Cancer of Unknown Primary, NOS | PLEURA | TRUE | FEMALE | NSCLC | PEMETREXED | 1 |
| 57 | Adenocarcinoma, NOS | LIVER | TRUE | MALE | PAAD | FOLFIRINOX | 1 |
| 58 | Cancer of Unknown Primary | LYMPH | TRUE | FEMALE | OVT | PACLITAXEL | 0 |
| 59 | Poorly Differentiated Carcinoma, NOS | SOFT_TISSUE | TRUE | FEMALE | NSCLC | PEMBROLIZUMAB/ PEMETREXED/CARBOPLATIN | 0 |
| 60 | Adenocarcinoma, NOS | LIVER | FALSE | MALE | GINET | GEMCITABINE/CISPLATIN | 1 |
| 61 | Adenocarcinoma, NOS | STOMACH | TRUE | MALE | EGC | OXALIPLATIN/FLUOROURACIL /LEUCOVORIN | 1 |
| 62 | Squamous Cell Carcinoma, NOS | BONE | FALSE | MALE | MEL | CETUXIMAB/CARBOPLATIN /FLUOROURACIL | 1 |
| 63 | Adenocarcinoma, NOS | LIVER | TRUE | FEMALE | PAAD | FOLFIRINOX | 1 |
| 64 | Poorly Differentiated Carcinoma, NOS | LYMPH | FALSE | MALE | EGC | PEMBROLIZUMAB | 1 |
| 65 | Adenocarcinoma, NOS | LIVER | FALSE | FEMALE | OVT | CAPECITABINE | 1 |
| 66 | Cancer of Unknown Primary | LIVER | TRUE | FEMALE | PAAD | FOLFIRINOX | 1 |
| 67 | Squamous Cell Carcinoma, NOS | LYMPH | TRUE | FEMALE | NSCLC | NIVOLUMAB | 1 |
| 68 | Neuroendocrine Tumor, NOS | LIVER | TRUE | FEMALE | PANET | OCTREOTIDE | 0 |
| 69 | Adenocarcinoma, NOS | LIVER | FALSE | FEMALE | EGC | GEMCITABINE/CISPLATIN | 1 |
| 70 | Squamous Cell Carcinoma, NOS | SOFT_TISSUE | FALSE | FEMALE | BLCA | PEMBROLIZUMAB | 1 |
| 71 | Cancer of Unknown Primary | LYMPH | TRUE | FEMALE | NSCLC | PEMETREXED/CISPLATIN | 0 |
| 72 | Adenocarcinoma, NOS | BRAIN | TRUE | FEMALE | COADREAD | OXALIPLATIN/FLUOROURACIL /LEUCOVORIN | 0 |

**Table S3 continued from previous page**

| | | | | | | | |
|---|---|---|---|---|---|---|---|
| 73 | Poorly Differentiated Carcinoma, NOS | LUNG | TRUE | FEMALE | OVT | CARBOPLATIN/DOXORUBICIN | 1 |
| 74 | Squamous Cell Carcinoma, NOS | LYMPH | SOFT FALSE | MALE | MEL | GEFITINIB | 1 |
| 75 | Adenocarcinoma, NOS | SOFT_TISSUE | TRUE | FEMALE | NSCLC | PEMETREXED/CARBOPLATIN | 1 |
| 76 | Adenocarcinoma, NOS | PERITONEUM | TRUE | MALE | EGC | OXALIPLATIN/FLUOROURACIL /LEUCOVORIN | 1 |
| 77 | Adenocarcinoma, NOS | SOFT_TISSUE | TRUE | FEMALE | OVT | CARBOPLATIN/PACLITAXEL | 1 |
| 78 | Adenocarcinoma, NOS | LIVER | TRUE | FEMALE | PAAD | FOLFIRINOX | 1 |
| 79 | Adenocarcinoma, NOS | LIVER | FALSE | FEMALE | NSCLC | GEMCITABINE/CISPLATIN | 1 |
| 80 | Cancer of Unknown Primary | LYMPH | FALSE | MALE | EGC | ETOPOSIDE/CARBOPLATIN | 1 |
| 81 | Adenocarcinoma, NOS | LUNG | TRUE | FEMALE | NSCLC | PEMETREXED/CARBOPLATIN | 0 |
| 82 | Cancer of Unknown Primary | UNKNOWN | FALSE | MALE | DIFG | PACLITAXEL/GEMCITABINE | 0 |
| 83 | Adenocarcinoma, NOS | SOFT_TISSUE | TRUE | MALE | PAAD | FOLFIRINOX | 1 |
| 84 | Adenocarcinoma, NOS | LIVER | TRUE | FEMALE | BRCA | CAPECITABINE | 0 |
| 85 | Cancer of Unknown Primary, NOS | LIVER | FALSE | MALE | PAAD | GEMCITABINE/CISPLATIN | 1 |
| 86 | Cancer of Unknown Primary | SOFT_TISSUE | FALSE | MALE | BLCA | OXALIPLATIN/FLUOROURACIL /LEUCOVORIN | 1 |
| 87 | Cancer of Unknown Primary, NOS | LIVER | FALSE | MALE | NSCLC | GEMCITABINE/CISPLATIN | 1 |
| 88 | Cancer of Unknown Primary, NOS | BOWEL | TRUE | MALE | EGC | OXALIPLATIN/FLUOROURACIL /LEUCOVORIN | 1 |
| 89 | Neuroendocrine Carcinoma, NOS | LIVER | FALSE | FEMALE | NSCLC | ETOPOSIDE/CISPLATIN | 0 |
| 90 | Neuroendocrine Tumor, NOS | BONE | FALSE | MALE | NSCLC | ETOPOSIDE/CARBOPLATIN | 1 |
| 91 | Adenocarcinoma, NOS | LIVER | FALSE | FEMALE | MEL | GEMCITABINE/CISPLATIN | 1 |
| 92 | Small Cell Carcinoma of Unknown Primary | LIVER | TRUE | MALE | PRAD | DOCETAXEL/CARBOPLATIN | 1 |

| | | | | | | | |
|---|---|---|---|---|---|---|---|
| 93 | Cancer of Unknown Primary | LIVER | FALSE | MALE | EGC | GEMCITABINE/CISPLATIN | 1 |
| 94 | Cancer of Unknown Primary | LIVER | TRUE | MALE | PAAD | OXALIPLATIN/FLUOROURACIL /LEUCOVORIN | 1 |
| 95 | Small Cell Carcinoma of Unknown Primary | SOFT_TISSUE | TRUE | MALE | EGC | OXALIPLATIN/FLUOROURACIL /LEUCOVORIN | 1 |
| 96 | Cancer of Unknown Primary | LYMPH | FALSE | MALE | MEL | CEMIPLIMAB | 0 |
| 97 | Cancer of Unknown Primary | LYMPH | FALSE | MALE | EGC | GEMCITABINE/CISPLATIN | 0 |
| 98 | Squamous Cell Carcinoma, NOS | SKIN | TRUE | MALE | HNSCC | NIVOLUMAB | 0 |
| 99 | Cancer of Unknown Primary | LIVER | FALSE | MALE | BRCA | CARBOPLATIN/PACLITAXEL | 1 |
| 100 | Adenocarcinoma, NOS | LIVER | TRUE | MALE | PAAD | OLAPARIB | 1 |
| 101 | Cancer of Unknown Primary | LIVER | FALSE | FEMALE | NSCLC | GEMCITABINE/CISPLATIN | 1 |
| 102 | Poorly Differentiated Carcinoma, NOS | LUNG | TRUE | MALE | NSCLC | PEMBROLIZUMAB | 0 |
| 103 | Adenocarcinoma, NOS | LUNG | TRUE | FEMALE | UCEC | CARBOPLATIN | 0 |
| 104 | Adenocarcinoma, NOS | LIVER | FALSE | FEMALE | UCEC | GEMCITABINE/CISPLATIN | 1 |
| 105 | Adenocarcinoma, NOS | LIVER | TRUE | MALE | COADREAD | FOLFIRI | 1 |
| 106 | Adenocarcinoma, NOS | LIVER | FALSE | FEMALE | BRCA | GEMCITABINE/CISPLATIN | 1 |
| 107 | Cancer of Unknown Primary, NOS | PLEURA | TRUE | FEMALE | BRCA | CAPECITABINE | 1 |
| 108 | Adenocarcinoma, NOS | BILIARY TRACT | FALSE | FEMALE | PAAD | GEMCITABINE/CISPLATIN | 1 |
| 109 | Adenocarcinoma, NOS | LIVER | TRUE | MALE | COADREAD | OXALIPLATIN/FLUOROURACIL /LEUCOVORIN | 0 |
| 110 | Cancer of Unknown Primary | OTHER | FALSE | FEMALE | OVT | ATEZOLIZUMAB/CARBOPLATIN /ETOPOSIDE | 1 |

| | | | | | | | |
|---|---|---|---|---|---|---|---|
| 111 | Cancer of Unknown Primary | LIVER | TRUE | MALE | CHOL | GEMCITABINE/CISPLATIN | 0 |
| 112 | Squamous Cell Carcinoma, NOS | VULVA | SOFT FALSE | FEMALE | NSCLC | BEVACIZUMAB/CARBOPLATIN /PACLITAXEL | 0 |
| 113 | Cancer of Unknown Primary | ADRENAL GLAND | FALSE | MALE | NSCLC | NIVOLUMAB | 1 |
| 114 | Neuroendocrine Tumor, NOS | BOWEL | SOFT FALSE | MALE | GINET | OXALIPLATIN/FLUOROURACIL /LEUCOVORIN | 0 |
| 115 | Neuroendocrine Carcinoma, NOS | LIVER | FALSE | MALE | EGC | ETOPOSIDE/CARBOPLATIN | 0 |
| 116 | Adenocarcinoma, NOS | OTHER | FALSE | MALE | RCC | GEMCITABINE/CISPLATIN | 0 |
| 117 | Adenocarcinoma, NOS | LYMPH | TRUE | FEMALE | NSCLC | PEMETREXED/CARBOPLATIN | 1 |
| 118 | Adenocarcinoma, NOS | LIVER | TRUE | MALE | EGC | OXALIPLATIN/FLUOROURACIL /LEUCOVORIN | 0 |
| 119 | Neuroendocrine Tumor, NOS | LIVER | TRUE | FEMALE | PANET | CAPECITABINE/TEMOZOLOMIDE | 0 |
| 120 | Cancer of Unknown Primary | CERVIX | FALSE | FEMALE | NSCLC | OXALIPLATIN/FLUOROURACIL /LEUCOVORIN | 0 |
| 121 | Poorly Differentiated Carcinoma, NOS | LYMPH | TRUE | MALE | MEL | PEMBROLIZUMAB | 0 |
| 122 | Poorly Differentiated Carcinoma, NOS | CERVIX | FALSE | FEMALE | NSCLC | CARBOPLATIN/PACLITAXEL | 1 |
| 123 | Adenocarcinoma, NOS | LIVER | FALSE | FEMALE | NSCLC | FOLFIRINOX | 0 |
| 124 | Cancer of Unknown Primary | THYROID | TRUE | MALE | HNSCC | PEMBROLIZUMAB | 1 |
| 125 | Adenocarcinoma, NOS | PERITONEUM | FALSE | FEMALE | BRCA | CARBOPLATIN/GEMCITABINE | 1 |
| 126 | Adenocarcinoma, NOS | LIVER | FALSE | MALE | NSCLC | PACLITAXEL/GEMCITABINE | 1 |
| 127 | Cancer of Unknown Primary | OVARY | FALSE | FEMALE | OVT | OXALIPLATIN/FLUOROURACIL /LEUCOVORIN | 1 |
| 128 | Neuroendocrine Carcinoma, NOS | LIVER | FALSE | MALE | DIFG | DOCETAXEL | 0 |
| 129 | Cancer of Unknown Primary | OTHER | FALSE | MALE | MEL | PEMETREXED | 1 |

| | | | | | | | |
|---|---|---|---|---|---|---|---|
| 130 | Poorly Differentiated Carcinoma, NOS | LIVER | FALSE | MALE | BLCA | ETOPOSIDE/CARBOPLATIN | 1 |
| 131 | Neuroendocrine Tumor, NOS | LIVER | TRUE | FEMALE | GINET | OCTREOTIDE | 0 |
| 132 | Adenocarcinoma, NOS | LIVER | TRUE | MALE | PAAD | PACLITAXEL/GEMCITABINE | 0 |
| 133 | Adenocarcinoma, NOS | LIVER | TRUE | MALE | EGC | OXALIPLATIN/FLUOROURACIL /LEUCOVORIN | 1 |
| 134 | Poorly Differentiated Carcinoma, NOS | LIVER | TRUE | MALE | CHOL | GEMCITABINE/CISPLATIN | 1 |
| 135 | Cancer of Unknown Primary, NOS | LYMPH | FALSE | FEMALE | BRCA | TOPOTECAN | 1 |
| 136 | Poorly Differentiated Carcinoma, NOS | LIVER | TRUE | MALE | BLCA | GEMCITABINE/CISPLATIN | 1 |
| 137 | Cancer of Unknown Primary | LUNG | FALSE | FEMALE | OVT | PEMBROLIZUMAB | 0 |
| 138 | Neuroendocrine Carcinoma, NOS | BRAIN | TRUE | FEMALE | NSCLC | CARBOPLATIN/PACLITAXEL | 0 |
| 139 | Cancer of Unknown Primary | LIVER | FALSE | FEMALE | DIFG | NIVOLUMAB | 0 |
| 140 | Poorly Differentiated Carcinoma, NOS | LUNG | TRUE | FEMALE | NSCLC | PEMETREXED/CARBOPLATIN | 0 |
| 141 | Poorly Differentiated Carcinoma, NOS | BOWEL | TRUE | FEMALE | OVT | CARBOPLATIN/GEMCITABINE | 1 |
| 142 | Poorly Differentiated Carcinoma, NOS | LUNG | TRUE | MALE | PRAD | DOCETAXEL | 0 |
| 143 | Poorly Differentiated Carcinoma, NOS | SOFT_TISSUE | TRUE | FEMALE | BRCA | CAPECITABINE | 0 |
| 144 | Adenocarcinoma, NOS | LIVER | FALSE | MALE | NSCLC | CABOZANTINIB | 1 |
| 145 | Adenocarcinoma, NOS | LIVER | TRUE | FEMALE | PAAD | FOLFIRINOX | 1 |

**Table S3 continued from previous page**

| | | | | | | | |
|---|---|---|---|---|---|---|---|
| 146 | Neuroendocrine Tumor, NOS | LIVER | TRUE | MALE | PANET | CAPECITABINE/TEMOZOLOMIDE | 0 |
| 147 | Cancer of Unknown Primary | LIVER | FALSE | FEMALE | COADREAD | GEMCITABINE/CISPLATIN | 1 |
| 148 | Adenocarcinoma, NOS | LIVER | TRUE | MALE | PAAD | FOLFIRINOX | 1 |
| 149 | Poorly Differentiated Carcinoma, NOS | HEAD_NECK | FALSE | MALE | GINET | CETUXIMAB/CARBOPLATIN /FLUOROURACIL | 1 |
| 150 | Neuroendocrine Tumor, NOS | LIVER | TRUE | MALE | PANET | EVEROLIMUS | 0 |
| 151 | Cancer of Unknown Primary, NOS | SOFT_TISSUE | TRUE | FEMALE | BLCA | BEVACIZUMAB/PACLITAXEL /CISPLATIN | 1 |
| 152 | Adenocarcinoma, NOS | PERITONEUM | TRUE | FEMALE | EGC | OXALIPLATIN/FLUOROURACIL /LEUCOVORIN | 1 |
| 153 | Adenocarcinoma, NOS | LIVER | TRUE | FEMALE | PAAD | PACLITAXEL/GEMCITABINE | 1 |
| 154 | Adenocarcinoma, NOS | LIVER | SOFT FALSE | FEMALE | BRCA | OLAPARIB | 1 |
| 155 | Adenocarcinoma, NOS | LIVER | TRUE | FEMALE | PAAD | FOLFIRINOX | 1 |
| 156 | Poorly Differentiated Carcinoma, NOS | BONE | TRUE | FEMALE | NSCLC | CARBOPLATIN/PACLITAXEL | 1 |
| 157 | Adenocarcinoma, NOS | LYMPH | TRUE | MALE | PAAD | FOLFIRINOX | 1 |

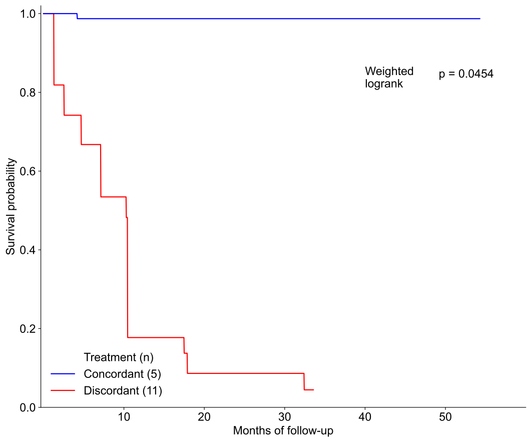


(a) Invasive Breast Carcinoma (BRCA)

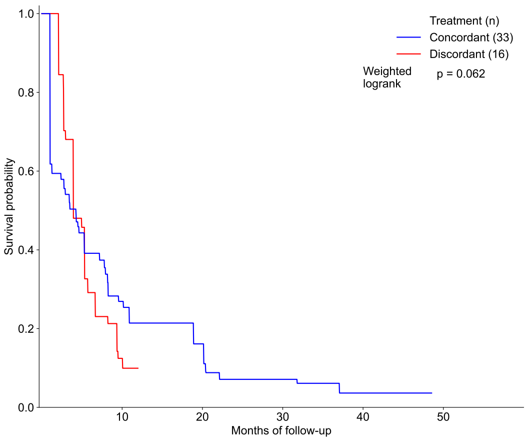


(b) Gastrointestinal (GI) group

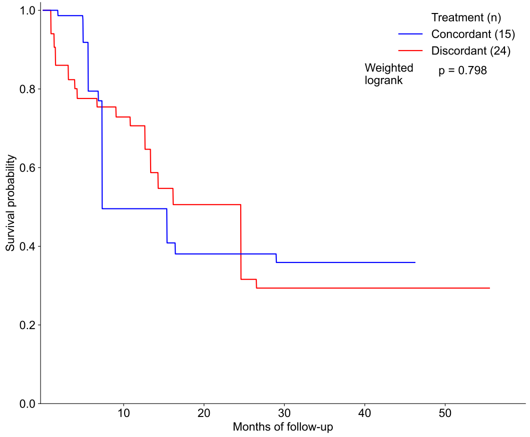


(c) Lung

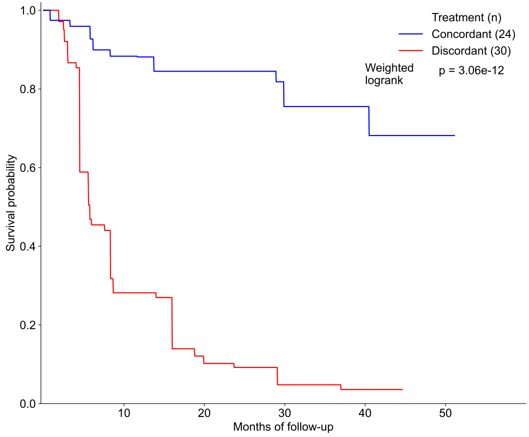


(d) Other OncoNPC cancer types

Figure S7

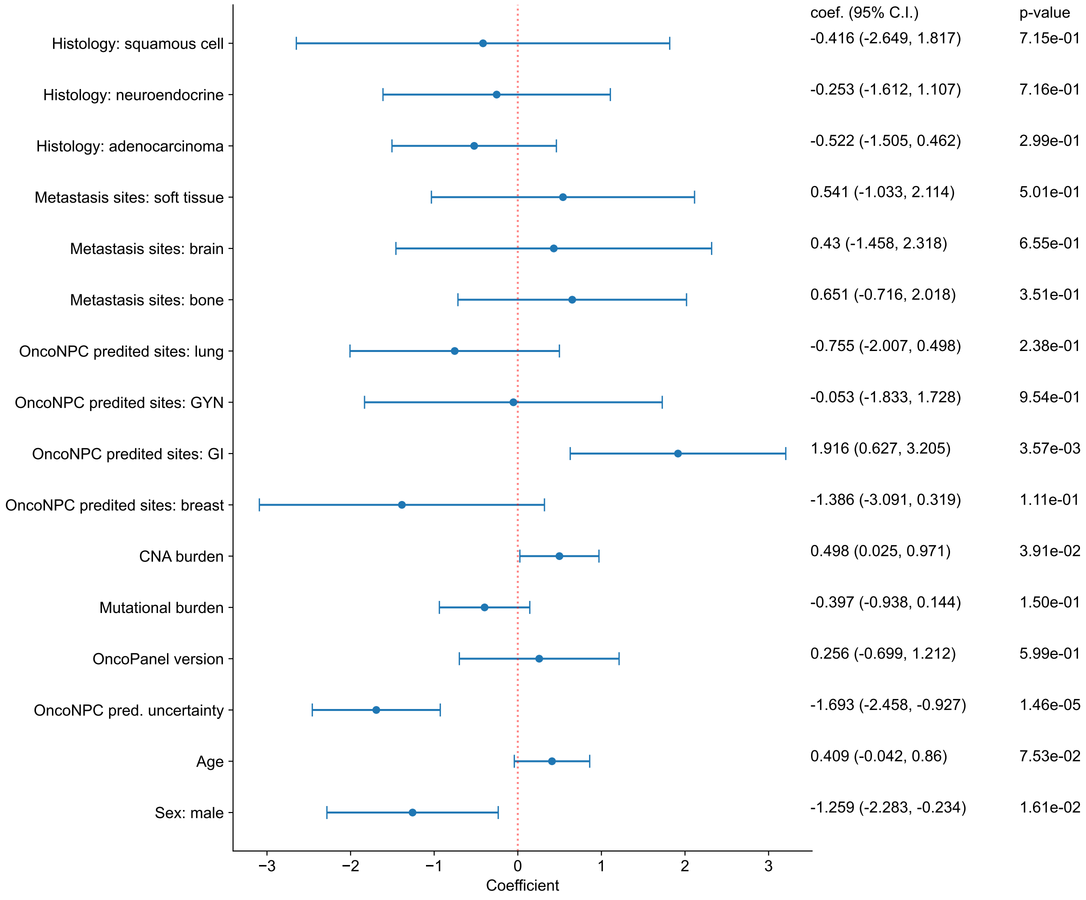
coef. (95% C.I.)
p-value
Histology: squamous cell
-0.416 (-2.649, 1.817)
7.15e-01
Histology: neuroendocrine
-0.253 (-1.612, 1.107)
7.16e-01
Histology: adenocarcinoma
-0.522 (-1.505, 0.462)
2.99e-01
Metastasis sites: soft tissue
0.541 (-1.033, 2.114)
5.01e-01
Metastasis sites: brain
0.43 (-1.458, 2.318)
6.55e-01
Metastasis sites: bone
0.651 (-0.716, 2.018)
3.51e-01
OncoNPC predited sites: lung
-0.755 (-2.007, 0.498)
2.38e-01
OncoNPC predited sites: GYN
-0.053 (-1.833, 1.728)
9.54e-01
OncoNPC predited sites: GI
1.916 (0.627, 3.205)
3.57e-03
OncoNPC predited sites: breast
-1.386 (-3.091, 0.319)
1.11e-01
CNA burden
0.498 (0.025, 0.971)
3.91e-02
Mutational burden
-0.397 (-0.938, 0.144)
1.50e-01
OncoPanel version
0.256 (-0.699, 1.212)
5.99e-01
OncoNPC pred. uncertainty
-1.693 (-2.458, -0.927)
1.46e-05
Age
0.409 (-0.042, 0.86)
7.53e-02
Sex: male
-1.259 (-2.283, -0.234)
1.61e-02
−3
−2
−1
0
1
2
3
Coefficient

Figure S8
